# Alzheimer’s Disease Sequencing Project Release 4 Whole Genome Sequencing Dataset

**DOI:** 10.1101/2024.12.03.24317000

**Authors:** Yuk Yee Leung, Wan-Ping Lee, Amanda B Kuzma, Heather Nicaretta, Otto Valladares, Prabhakaran Gangadharan, Liming Qu, Yi Zhao, Youli Ren, Po-Liang Cheng, Pavel P Kuksa, Hui Wang, Heather White, Zivadin Katanic, Lauren Bass, Naveen Saravanan, Emily Greenfest-Allen, Maureen Kirsch, Laura Cantwell, Taha Iqbal, Nicholas R Wheeler, John J. Farrell, Congcong Zhu, Shannon L Turner, Tamil I Gunasekaran, Pedro R Mena, Jimmy Jin, Luke Carter, Alzheimer’s Disease Sequencing Project, Xiaoling Zhang, Badri N Vardarajan, Arthur Toga, Michael Cuccaro, Timothy J Hohman, William S Bush, Adam C Naj, Eden Martin, Clifton Dalgard, Brian W Kunkle, Lindsay A Farrer, Richard P Mayeux, Jonathan L Haines, Margaret A Pericak-Vance, Gerard D Schellenberg, Li-San Wang

## Abstract

The Alzheimer’s Disease Sequencing Project (ADSP) is a national initiative to understand the genetic architecture of Alzheimer’s Disease and Related Dementias (AD/ADRD) by sequencing whole genomes of affected participants and age-matched cognitive controls from diverse populations. The Genome Center for Alzheimer’s Disease (GCAD) processed whole-genome sequencing data from 36,361 ADSP participants, including 35,014 genetically unique participants of which 45% are from non-European ancestry, across 17 cohorts in 14 countries in this fourth release (R4). This sequencing effort identified 387 million bi-allelic variants, 42 million short insertions/deletions, and 2.2 million structural variants. Annotations and quality control data are available for all variants and samples. Additionally, detailed phenotypes from 15,927 participants across 10 domains are also provided. A linkage disequilibrium panel was created using unrelated AD cases and controls. Researchers can access and analyze the genetic data via NIAGADS Data Sharing Service, the VariXam tool, or NIAGADS GenomicsDB.

## Introduction

Alzheimer’s disease (AD) is a neurodegenerative condition characterized by the abnormal buildup of amyloid-β peptides in extracellular plaques and hyperphosphorylated tau in intracellular neurofibrillary tangles. This progressive neurodegeneration results in a gradual decline in cognitive and functional abilities. Genetic variants play a significant role in the development of late-onset AD (LOAD). The first notable finding in 1993 reported the *ε*4 allele of the apolipoprotein E (*APOE*) gene was associated with the risk of developing AD [1]. The identification of additional genetic factors for LOAD accelerated with the advent of high-throughput genomic technologies, such as genotype arrays, in the late 2000s [2, 3]. Since then, the list of genetic factors continues to expand with the inclusion of bigger samples sizes through international consortium efforts, notably between the Alzheimer’s Disease Genetics Consortium (ADGC) and International Genomics of Alzheimer’s Project (IGAP) [4–6]. However, most of these identified genetic variants are common alleles with individually small causal effects on disease susceptibility as the studies focused on genotype array data. These variants contribute minimally to the overall genetic liability for the disease, as a study showed that the single nucleotide polymorphism (SNP)-heritability estimate from the largest AD genome-wide association studies (GWAS) to date is 3.1% [7], which is significantly smaller than the heritability estimates for AD obtained from twin studies, which ranges from 60 to 80% [8, 9].

Whole-genome sequencing (WGS) can address this missing heritability challenge by 1) providing a more comprehensive view of the genetic architecture via a full-spectrum of variants, and 2) identifying rare variants with potentially larger phenotypic effects. Analyzing many samples is necessary to address the above gaps. WGS studies are more costly compared to genotyping array, and as a result fewer participants of non-European descent have been sequenced [10]. Expanding AD research to ancestrally diverse populations is crucial for several reasons. Most genetic studies of Alzheimer’s disease (AD) have focused on non-Hispanic White (NHW) populations. However, genetic risk factors identified in NHW populations may not fully explain the observed ethnic disparities in AD. For instance, while APOE ε4 is a significant predictor of late-onset AD in NHW individuals, its predictive power is weaker and more inconsistent in African American (AA) and Hispanic or Latino (HL) populations [11–13]. Investigating how genetic risk factors for AD vary among ethnic groups could pave the way for more effective, tailored treatments and interventions. Notably, certain genetic variants—such as those in *SORL1*, *ABCA7*, and *ACE*—exhibit stronger associations with AD risk in specific groups, including Asians [14], AA [15], and Israeli-Arabs [16]. These findings suggest that gene therapies targeting these genes may have varying levels of effectiveness across different ethnicities.

Funded through cooperative agreements and research grants, the Alzheimer’s Disease Sequencing Project (ADSP) brings together 497 investigators from institutions worldwide. In 2023, it successfully completed the “Follow-Up Study (FUS) Phase” (the third phase of ADSP), sequencing existing cohorts of AA and pan-Hispanic ancestry at The American Genome Center at the Uniformed Services University of the Health Sciences (USUHS) and John P. Hussman Institute for Human Genomics (HIHG). This effort was conducted in collaboration with established NIH-funded AD infrastructure like the National Cell Repository for Alzheimer’s Disease (NCRAD), National Institute on Aging Genetics of Alzheimer’s Disease Data Storage Site (NIAGADS), and the Genome Center for Alzheimer’s Disease (GCAD). Additionally, participants from NHW and Asian ancestries have been sequenced. All genomes, including those from previous ADSP phases and other collaborative projects, have been processed using a unified pipeline, subjected to comprehensive quality control, and annotated using various resources. This collection, the Release 4 (R4) of ADSP data, forms the world’s largest publicly available AD genome resource.

## Results

### Sequence existing ancestrally diverse cohorts via the ADSP Follow-Up Study (ADSP-FUS)

The ADSP-FUS is a National Institute on Aging (NIA) initiative focused on identifying genetic risk and protective variants for AD by expanding the ADSP cohorts beyond primarily participants with NHW. Given limitations in population diversity in the ADSP, the ADSP-FUS was designed to sequence existing ancestrally diverse and unique cohorts. ADSP-FUS 2.0 (The Diverse Population Initiative) focuses on HL, non-Hispanic Black with African Ancestry, and Asian populations (e.g., the Asian cohort for Alzheimer’s disease). ADSP-FUS intends to sequence over 100,000 participants from diverse ancestries (**Methods** - **Sequence existing ancestrally diverse cohorts via the ADSP Follow-Up Study (FUS)**). ADSP has developed a workflow (**Supplementary Figure 1**) to support biospecimens processing, DNA preparation, and sequencing at USUHS and HIHG.

### Sample characteristics

20,771 WGS data (8,159 new) from the ADSP-FUS phases are included in the ADSP Release 4 (R4) dataset, bringing the total number to 36,361 across 17 cohorts/studies in 14 countries (**Figure 1A**). Sequencing was carried out at ten sequencing centers using Illumina technology. Most were generated using the PCR-free protocol (91%) and 150bp in read length (94%). Three sequencing platforms were used: Illumina 2000/2500 (7%), HiSeqX (35%), or NovaSeq (58%) machines. **Supplementary Table S1** provides more details on sequencing configurations. GCAD processed all 36,361 WGS samples from read mapping to variant calling using a standard pipeline (VCPA1.1) to harmonize all data and minimize batch effects [17] (**Methods** - **Dataflow and Sample processing protocol on SNVs and indels**).

**Figure 1.**
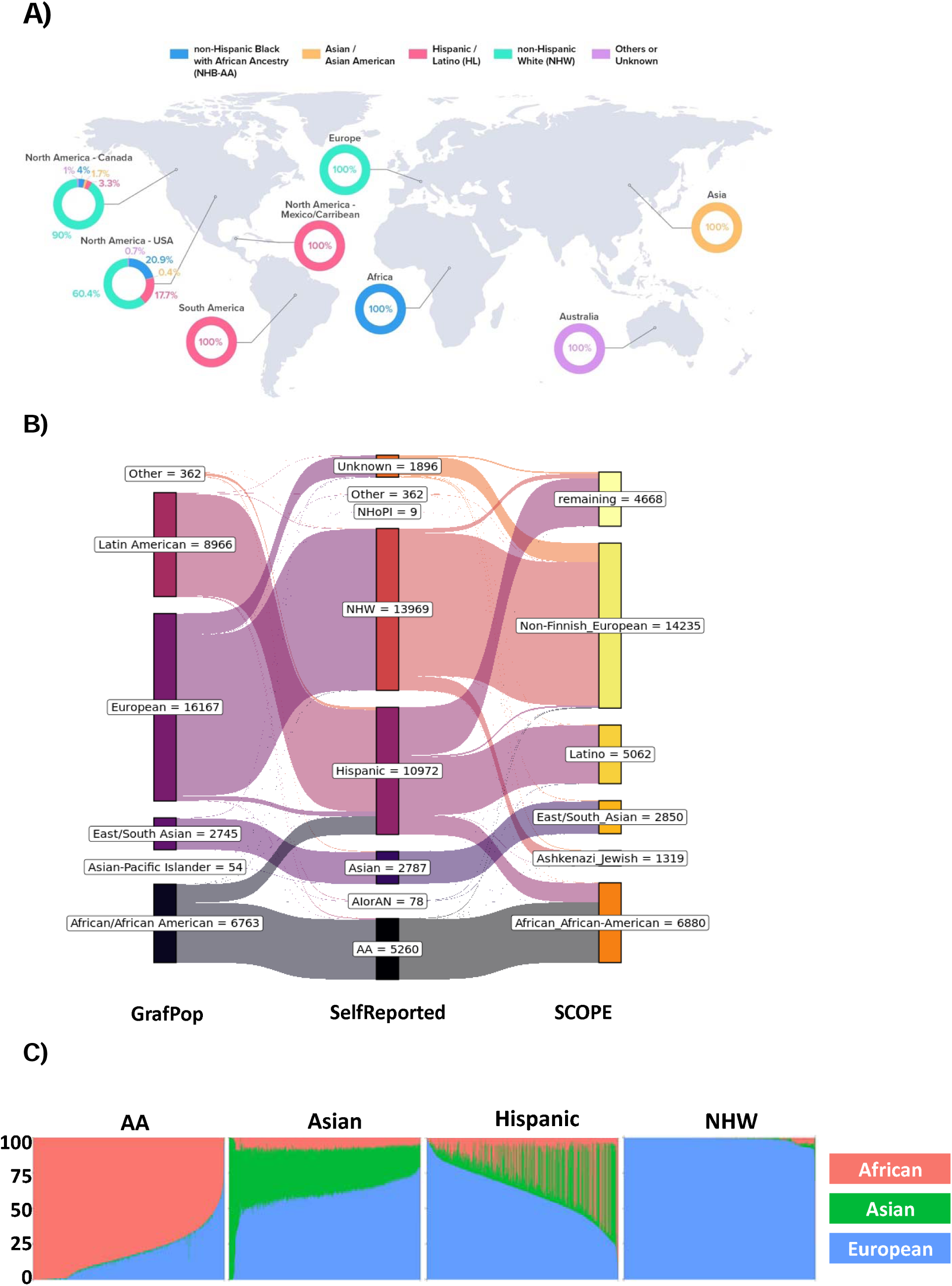
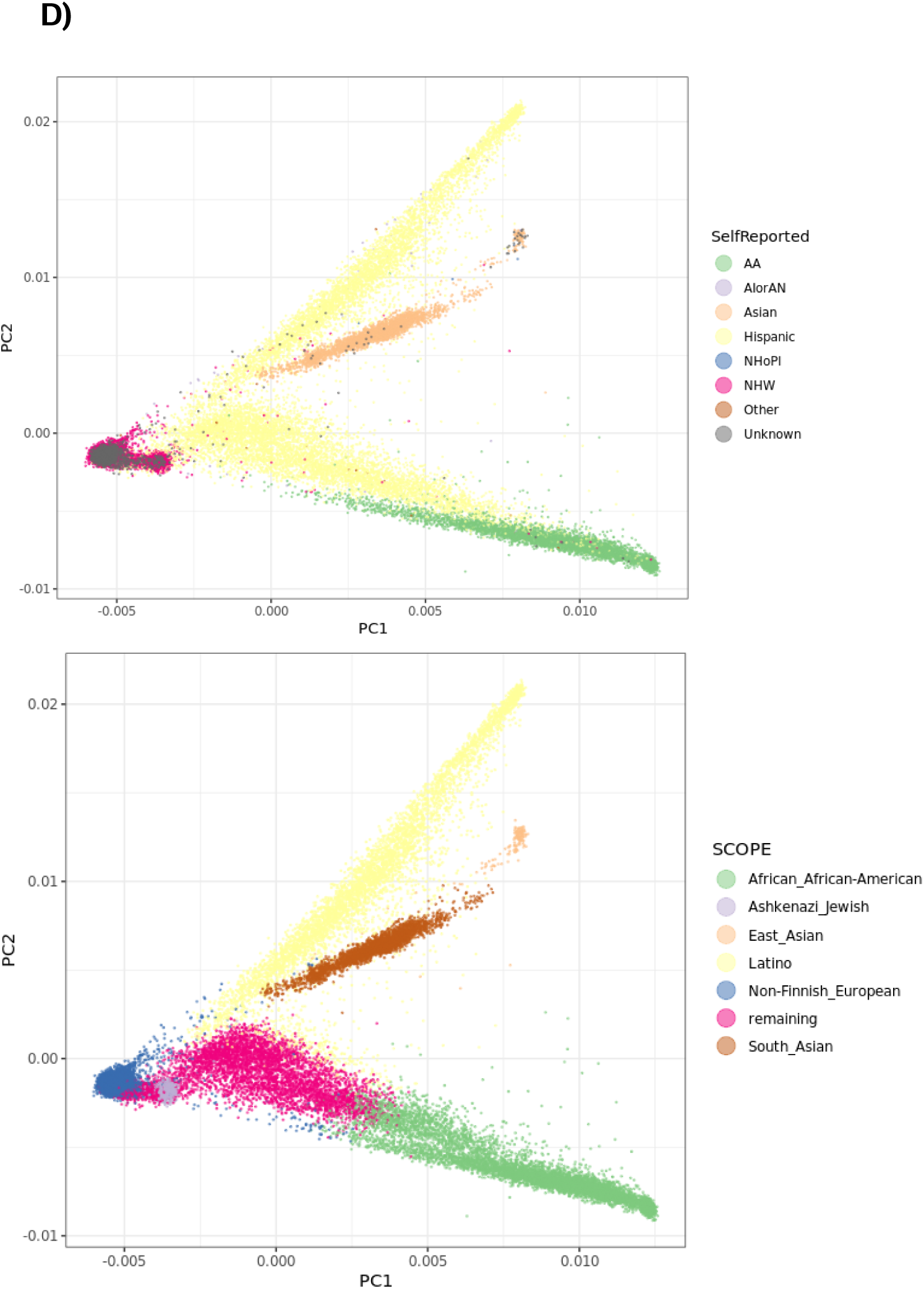
Participants in ADSP R4 dataset. **A)** Worldwide cohorts assembled for this ADSP R4 dataset. Non-Hispanic Black with African Ancestry (NHB-AA) samples are from Africa, and North America (Canada, USA); Asian and Asian American are from Asia, and North America (Canada, USA); Hispanic/Latino (HL) are from North America (Mexico/Caribbean, Canada, USA), and South America; non-Hispanic white (NHW) are from Europe and North America (Canada, USA). Lastly, some samples categorized as others or unknown and they are from Australia. **B)** Comparison of reported ethnicity against those inferred by GRAF-POP and SCOPE based methods. **C)** Estimated GRAF-pop ancestral components Pe, Pf, and Pa for all participants. **D)** PCA plot on R4 participants colored by reported ethnicity (top) or SCOPE.

Of these 36,361 samples, 35,014 participants are genetically unique. Based on ethnicity reported by cohorts, there are 5,260 AA, 78 American Indian or Alaskan Native (AIorAN), 2,787 Asian, 10,972 Hispanics, 9 Native Hawaiian or Pacific Islander (NHoPI), 13,969 NHW participants, and 1,896 participants were of unknown ethnicity. A breakdown of demographic information is summarized in **Figure 1A** and **Supplementary Table S2**.

We inferred the genetic ancestry of each individual using called genetic data to investigate the discordance between the reported and genetically inferred ancestries. Most discordance reported were in admixed participants [18], presenting additional challenges in identifying ancestry-specific variants. We used GRAF-pop [19] (**Methods - GRAF-POP**), which assumes that each individual is an admixture of three ancestral groups: European (e), African (f), and Asian (a). GRAF-pop estimates ancestry components *Pe*, *Pf*, and *Pa*, which are then used to assign participants to population groups, including European, African/African American, Latin American, Asian-Pacific Islander, and East/South Asian. Using the software’s default settings, the match rates between reported and genetically inferred ancestries were 99.4% for European, 98.3% for African American, 96.8% for Asian, and 80.0% for Hispanic participants (**Figures 1B and 1C**).

We also performed principal component analysis (PCA) based on genotypes derived from WGS (**Methods – Population substructure**). We selected common variants (MAF>0.02) of high quality and performed linkage disequilibrium (LD) pruning to yield 146,964 variants, then calculated principal components (PCs) and genetic relationship matrix (GRM). We then performed the ancestry inference analysis using 145,278 variants common in both the ADSP R4 and gnomAD data [20], a publicly available population genetics resource generated on 76,215 diverse samples. Subjects were assigned to an ancestry group in which it has the highest ancestry proportion value. The match rates between reported and genetically inferred ancestries by this method were 88.3% for Non-Finnish European and 8.7% for Ashkenazi Jewish, 98.7% for AA, 99.7% for Asian, and among Hispanic participants, 45.5% for Latino, 15% for AA and 37.8% for remaining. The remaining subjects are most likely from Caribbean region (**Figures 1B and 1D**).

### WGS sample quality assessment

We performed QC checks on all samples and 734 samples with quality issues **(Method – ADSP Sample level QC protocol**) and reassessed the quality of the callset (N=36,361). The mean read depth across samples is 40.4x with 99% of samples having a coverage >30x (**Figure 2A**). The per genome percentage of bases with the quality score greater than Q30 (sequencing error rate less than 0.1%) is 90.18±2.43%. On average, 98.92±2.13% reads of samples are mapped, and 94.14±2.76% of paired-end reads have both ends mapped.

**Figure 2.**
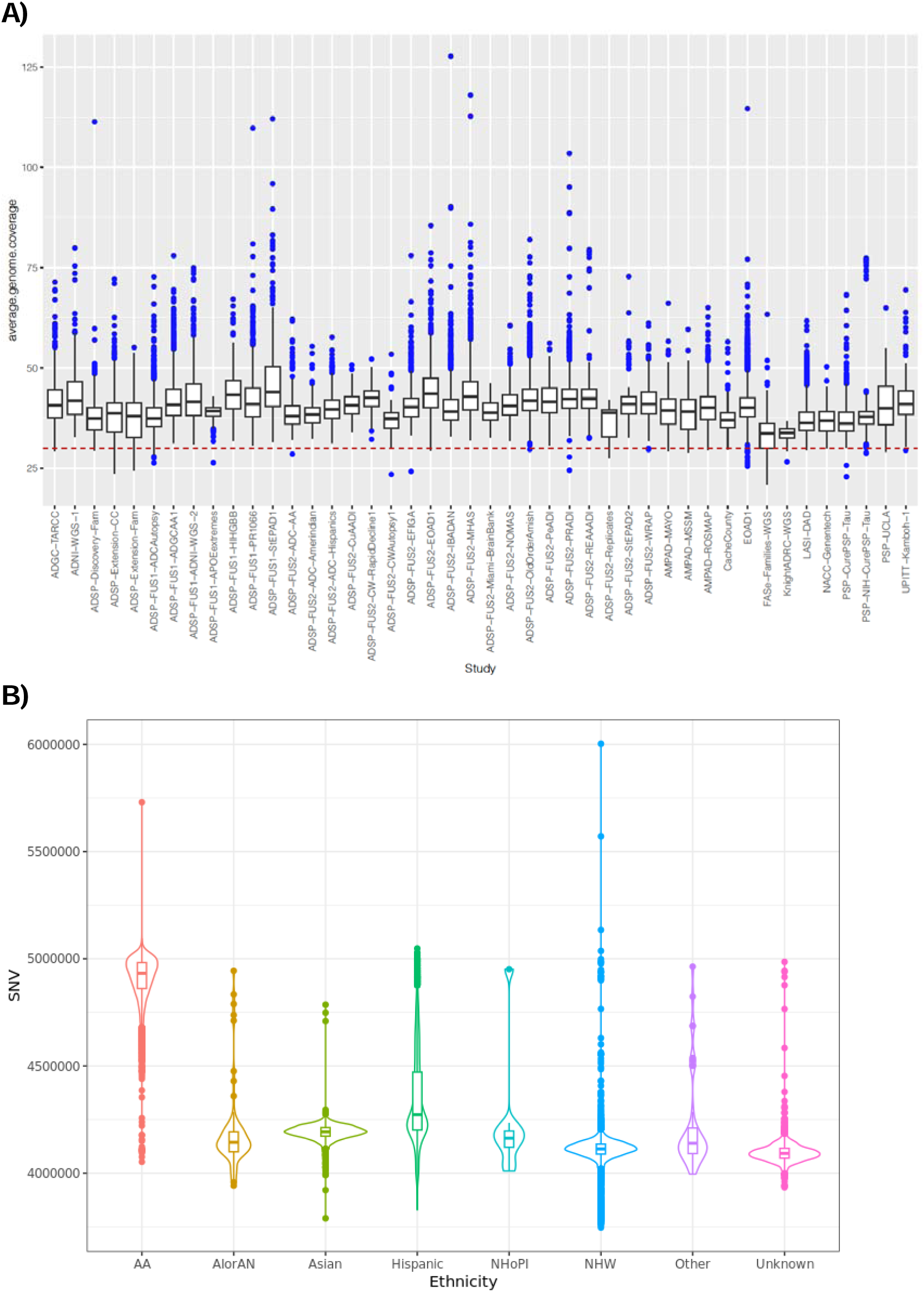
WGS sample quality. **A)** Coverage (30x) for the ADSP R4 data. Red dotted line indicates coverage value at 30. 99% of samples pass this threshold. **B)** Number of SNVs called per sample in each reported ethnic group. Line in each displayed boxplot denotes the mean value where each dot is a sample.

On average, each sample contained 4.3 million single nucleotide variants (SNVs) and 999,000 short insertions and deletions (indels). AA samples have the highest number of variants (4.9 million) followed by Hispanic samples (4.3 million), Asian samples (4.2 million), and NHW samples (4.1 million) (**Figures 2B, Supplementary Figure 2a**).

We assessed if the number of called variants were affected by sequencing configurations such as sequencing platform and use of PCR. We found that samples (regardless of ethnicities) sequenced by the PCRFree protocol tend to yield more variants, with the combination of NovaSeq + PCRFree returning the highest (**Supplementary Figure 2b,2c**).

### Bi-allelic variants (SNVs and indels) in ADSP R4

We called genotypes for all observed variants across all 36,361 samples and split the joint-called results into two VCF file sets. One VCF was generated using GATK4.1.1 on all SNVs and short indels. We identified 322,757,476 bi-allelic SNVs and 24,005,724 bi-allelic indels on autosomes, comprising 83% of the original GATK output, which will be the focus of this article. R4 data also contains 54,425,255 multi-allelic SNVs and indels on autosomes. A variant passes QC if it has a GATK “FILTER” = PASS or is in tranche >= 99.8%, DP>10 and GQ>20, call Rate>=80%, and is supported by <500 reads (**Method - ADSP Variant QC protocol**). Average call rate of the variants is high (97.0%). Details for other quality of the variants can be found in **Supplementary Table S3**.

The ADSP quality control (QC) protocol flagged 92.94% of autosomal variants, 299,620,924 SNVs and 22,674,845 indels, as high quality. Of the four major ethnicities, AA (N=5,260), Asian (N=2,787), Hispanic (N=10,972), and NHW (N=13,969), there are 101,227,106 (94,371,761 SNVs and 6,855,642 indels) for AA, 69,338,361 (64,882,876 SNVs and 4,455,485 indels) for Asian, 132,424,746 (123,631,863 SNVs and 8,792,883 indels) for Hispanic, and 135,672,855 (126,113,083 SNVs and 9,559,772 indels) for NHW (**Table 1, top**) respectively.

**Table 1.** – Number of variants (SNVs and indels) identified in the four major ethnic groups in ADSP R4 data, broken down allele frequency (AF) Shown in **top** is the variant count and percentages per ethnicity. Instead of the total number of variants identified, we showed at the **bottom** the ethnic specific variants.

| All | AA |  | Asian |  | Hispanic |  | NHW |  |
| --- | --- | --- | --- | --- | --- | --- | --- | --- |
| AF | Count | % | Count | % | Count | % | Count | % |
| Singleton | 48,980,233 | 48.4 | 38,123,455 | 55.0 | 61,467,493 | 46.4 | 78,473,265 | 57.8 |
| <0.1% | 27,252,406 | 26.9 | 15,126,076 | 21.8 | 48,762,383 | 36.8 | 43,759,910 | 32.3 |
| 0.1-1% | 11,674,387 | 11.5 | 8,038,369 | 11.6 | 12,218,350 | 9.2 | 5,653,864 | 4.2 |
| 1-5% | 5,806,358 | 5.7 | 2,370,814 | 3.4 | 4,054,972 | 3.1 | 2,242,076 | 1.7 |
| >5% | 7,514,019 | 7.4 | 5,679,746 | 8.2 | 5,921,548 | 4.5 | 5,543,740 | 4.1 |

| Unique | AA |  | Asian |  | Hispanic |  | NHW |  |
| --- | --- | --- | --- | --- | --- | --- | --- | --- |
| AF | Count | % | Count | % | Count | % | Count | % |
| Singleton | 33,348,856 | 76.5 | 29,033,386 | 70.1 | 44,486,085 | 67.5 | 61,301,483 | 76.0 |
| <0.1% | 10,107,022 | 23.2 | 9,289,546 | 22.4 | 20,809,053 | 31.6 | 19,206,135 | 23.8 |
| 0.1-1% | 151,690 | 0.3 | 2,916,818 | 7.0 | 644,326 | 1.0 | 133,697 | 0.2 |
| 1-5% | - | - | 157,348 | 0.4 | 1,967 | 0.0 | - | 0.0 |
| >5% | - | - | 787 | 0.0 | 1 | 0.0 | - | 0.0 |

Regarding allele frequency (AF), 52.53% of variants are singletons, followed by 40.62% rare variants with AF < 0.1%, 3.72% with AF 0.1-1%, 1.29% with AF 1-5%, and 1.84% with AF > 5%. The distribution of variants across AF ranges is consistent across ethnicities, with singletons comprising close to or more than 50% of the variants. AA, Asian, and Hispanic groups (13.16%, 11.61%, and 7.53%, respectively) have a higher proportion of variants with AF > 1% compared to the overall dataset and the NHW group (5.74% and 3.13%, respectively) (**Table 1, top**).

In terms of ethnic specific variants, there are 43,607,568 (40,417,070 SNVs and 3,190,498 indels), 41,397,885 (38,544,025 SNVs and 2,853,860 indels), 65,941,432 (61,362,910 SNVs and 4,578,522 indels), 80,641,315 (7,4475,786 SNVs and 6,165,529 indels) variants for AA, Asian, Hispanic, and NHW. Most of the ancestral specific variants are rare variants (AF < 0.1%, **Supplementary Table S4**) (**Table 1, bottom**).

We compared the ADSP R4 bi-allelic variants to gnomAD [21] (**Method – Comparison of gnomAD**). Of the ADSP R4 variants, 62.39% of SNVs and 57.33% of indels are reported in gnomAD. In terms of allele frequencies, 99.89% of variants with AF > 5%, 99.80% with 1% < AF ≤ 5%, 99.74% with 0.1% < AF ≤ 1%, 82.76% with AF ≤ 0.1%, and 41.08% of singletons are present in the gnomAD database.

### Annotation of genetic variants

The official ADSP annotation pipeline [22] was used to annotate all 347 million variants (SNVs and indels) (**Methods – Variant annotation protocol**, **Figure 3A**). Functional impact of variants was accessed using snpEff [23] (**Methods – LOF analyses**). We identified 224,594 high-impact loss-of-function variants: frameshift (39%), stop gained (27%), splice donor (16%), splice acceptor (12%), start lost (4%), and stop lost (2%) across 22,710 genes (**Figure 3B**). Among these, 1,295 genes were found to be intolerant to protein-truncating loss-of-function variants, indicated by a Loss Intolerance Probability (pLI) score of 1, suggesting the critical importance of these genes. We also provide annotation for all bi-allelic variants using FAVOR (**Methods – FAVOR annotation protocol**). 27.45 million variants of such are with CADD (phred score) of 20 or above, with over 63.64 million variants lying in super-enhancer regions.

**Figure 3.**
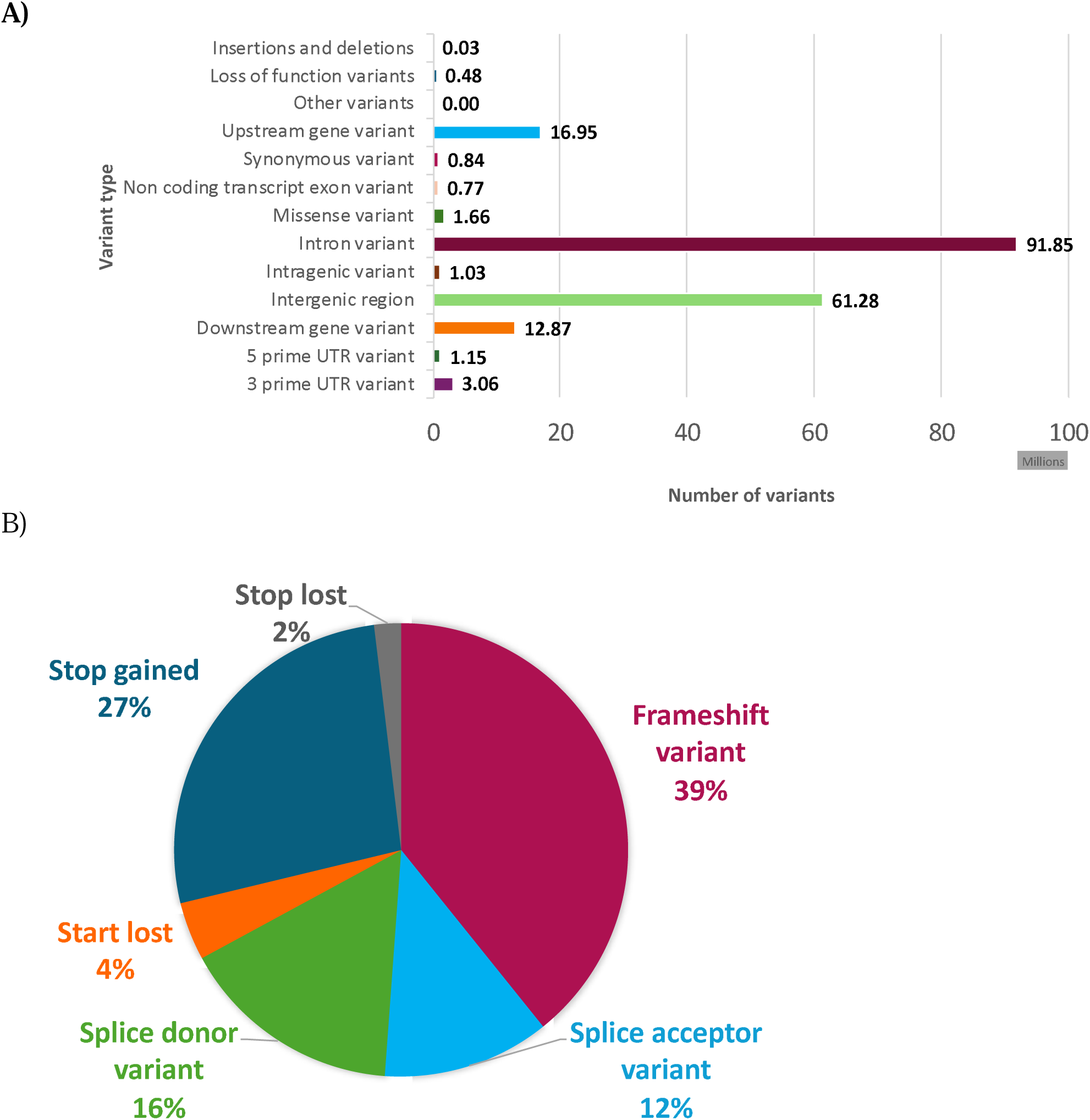
The distribution of variant types across the genome, with a specific focus on high-risk loss-of-function variants. **A)** Bar chart depicting the breakdown of the total number of variants across the genome, categorized by genomic annotation as follows: insertions and deletions, loss-of-function variants, upstream gene variants, synonymous variants, non-coding transcript exon variants, missense variants, intron variants, intragenic variants, intergenic variants, downstream variants, 5 prime UTR variants, and 3 prime UTR variants. **B)** The distribution of 224,594 loss-of-function variants is further broken down into the following categories: frameshift (39%), stop gained (27%), splice donor (16%), splice acceptor (12%), start lost (4%), and stop lost (2%).

### Structural variants in ADSP R4 samples

We applied the same protocol developed for the ADSP R3 dataset [24] to the R4 dataset **(Methods – Structural variant calling protocol)**. Individual Manta and Smoove callsets were initially merged for each sample, and then all samples were combined using SVIMMER (v0.1). GraphTyper (v2.7) was subsequently applied to the merged VCF for structural variant (SV) joint genotyping. Notably, only SVs larger than 10 Mbp were filtered. The final callset consists of a total of 6,796,267 SVs, including 4,101,354 deletions, 726,560 duplications, 558,860 insertions, and 1,409,493 inversions. Since an SV can be associated with multiple joint genotyping models, such as AGGREGATED, BREAKPOINT, BREAKPOINT1, BREAKPOINT2, and COVERAGE in GraphTyper2, some SVs appeared multiple times in the R4 SV VCF. After consolidating these models, the total number of unique SVs was reduced to 2,208,044, comprising 1,367,118 deletions, 184,367 duplications, 186,290 insertions, and 470,269 inversions.

On average, 15,640 high-quality SVs were identified per sample, including 7,813 deletions, 1,574 duplications, 6,246 insertions, and 7 inversions. Similar to the patterns observed with SNVs and indels, AA samples exhibited a higher number of SVs compared to other groups (**Figure 4**).

**Figure 4.**
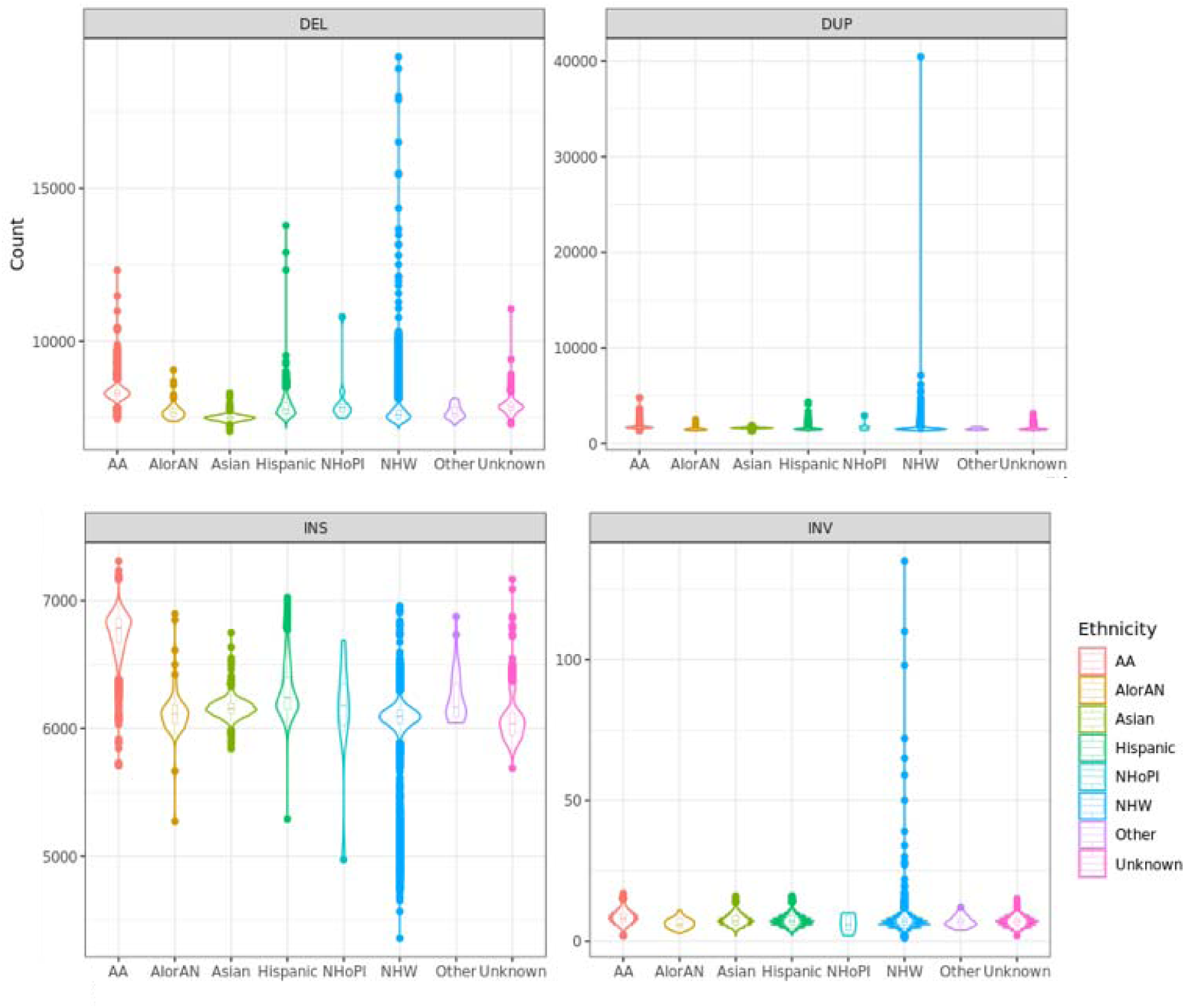
– Comparison of the number of SVs called in the ADSP R4 dataset across different reported ethnicities. SVs can be categorized into four different types of SVs: deletion (DEL), duplication (DUP), inversion (INV), and insertion (INS).

### LD reference panel from ADSP R4 data

Starting with the ADSP Integrated Phenotypes list which includes 32,236 samples (5,096 AA, 2,777 Asian, 10,438 Hispanic, 12,692 NHW, and 1,233 others), we constructed panels for AA, Asian, Hispanic, and NHW separately. The panels were built using both SNVs and indels. The number of variants included in each panel was as follows: 46,462,895 variants (43,494,096 SNVs and 2,968,799 indels) for AA, 25,779,737 variants (24,235,526 SNVs and 1,544,211 indels) for Asians, 57,683,258 variants (54,000,742 SNVs and 3,682,516 indels) for Hispanics, and 44,629,226 variants (41,794,882 SNVs and 2,834,344 indels) for NHW.

We performed emeraLD [25] with the following parameters, --mac > 5, --threshold 0.2, and -- window 5000000. Each segment was analyzed by 5Mb window with a 3Mb overlapping, then we concatenated all segments, removing duplicate records. As a result, we identified 3,153,513,864, 1,795,829,862, 4,990,587,680, and 3,205,008,552 pairs of variants for the AA, Asian, Hispanic, and NHW, respectively. Among these, the proportion of pairs with R^2^ > 0.8 was 6.3% for AA, 11.0% for Asians, 5.7% for Hispanics, and 9.2% for NHW. For R^2^ > 0.2, the proportions were 33.5%, 45.4%, 32.4%, and 40.9%, respectively.

### Harmonized phenotypic data for 28,000+ participants with WGS

The ADSP Phenotype Harmonization Consortium (ADSP-PHC) was established to unify detailed endophenotype data from various cohort studies. The group collaborates with ADSP to ensure high-quality phenotype harmonization across multiple domains, and document data availability and harmonization processes. Currently, available phenotypes from the ADSP-PHC include autopsy measures of neuropathology, fluid biomarkers of AD neuropathology, positron emission tomography measures of amyloid and tau pathology, structural brain imaging using magnetic resonance imaging, diffusion tensor imaging, longitudinal measures of cognition, and cardiovascular risk factor data. Harmonization methods are detailed in the **Methods** - **ADSP Phenotype Harmonization Consortium (ADSP-PHC)** section.

In order to ensure the highest quality harmonization is conducted, the ADSP-PHC harmonizes all available phenotypic data, regardless of sequencing status, which the research community can access directly from each cohort. ADSP-PHC deliverables are then subset to participants with available sequencing data. To increase the value and usage of the ADSP dataset, the ADSP-PHC has selected >9k phenotypes across 10 domains, expanding on the three domains released previously (NIAGADS ng00067.v9). **Figure 5** summarizes the harmonized data availability for more than 15,927 participants with whole genome sequencing and harmonized phenotypic data in NIAGADS. A data availability and explorer tool are available online via https://vmacdata.org/adsp-phc.

**Figure 5.**
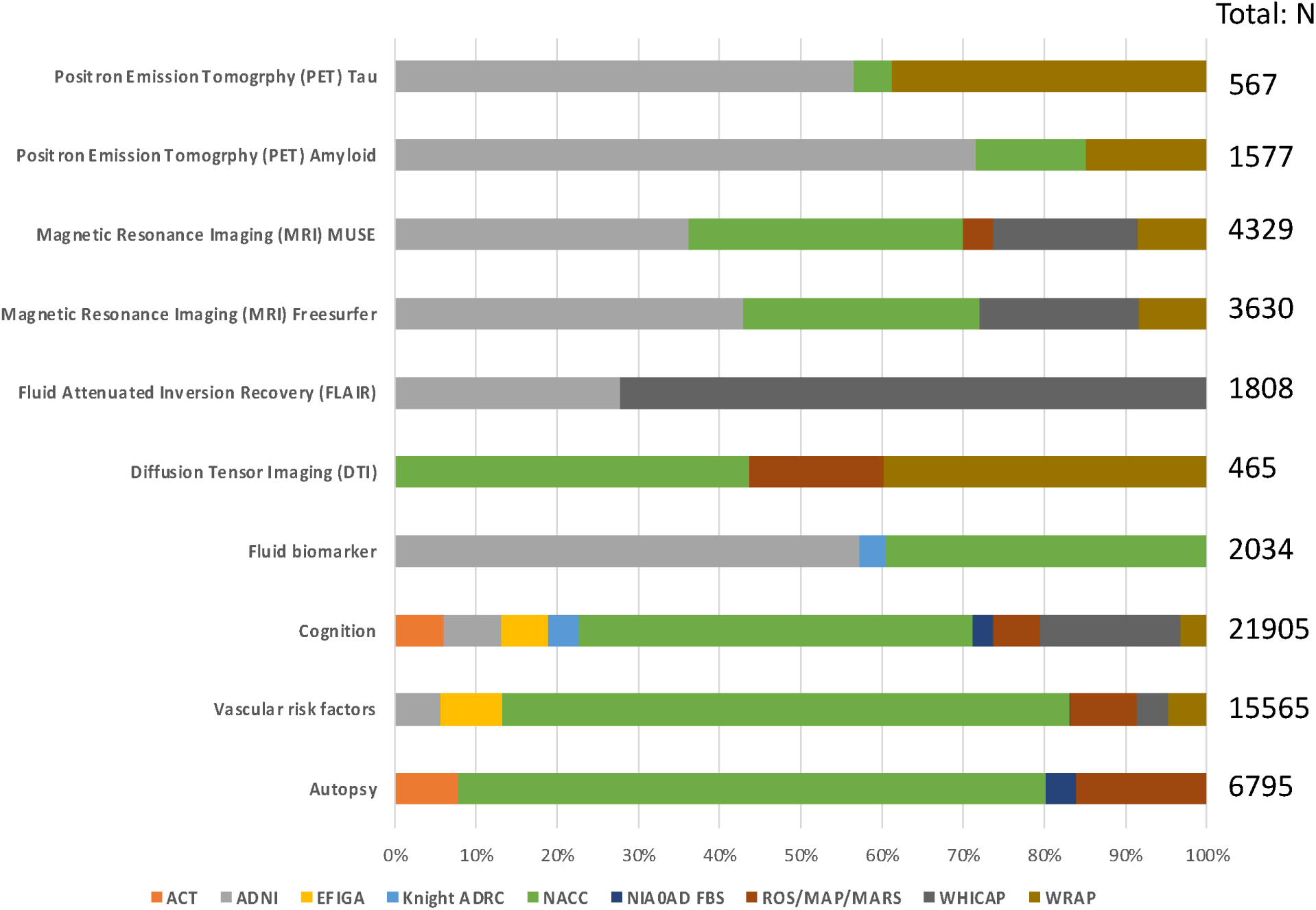
–ADSP-PHC Release (ng00067.v11) Sample sizes (“N” on the right) reflect individuals with ADSP sequencing data in R4.

### Summary of data files shared in this collection

All the described R4 data have been released in NIAGADS Data Sharing Service (DSS, https://dss.niagads.org/). These include individual level CRAMs, gVCFs, and SV VCFs, as well as aggregated files including the joint-genotyped VCFs from 35,014 unique individuals. We offer alternative solutions for users who may not require VCFs with detailed information for their analyses. These options include genotype information, full quality metrics, or ADSP QC details, and are organized by bi-allelic and multi-allelic variants. Additionally, some of these files are available in the CoreArray Genomic Data Structure (GDS) [26], an alternative format to VCF designed specifically for R users. Besides we provided sequencing methods, quality data metrics, variant metrics, phenotypes and readmes along with these data files. Annotation and LD reference panel files are available in open access. We summarize these files by features and file size in **Table 2**.

**Table 2.** – ADSP R4 released file set. All files are available under https://dss.niagads.org/datasets/ng00067/. Both individual (CRAMs, gVCFs, SV VCFs and phenotypes) and summary level files (VCFs, GDS, annotation files) are available. Annotation and LD reference panel files are also available in NIAGADS Open Access Data Portal https://dss.niagads.org/open-access-data-portal/.

| Descriptions |  | Genotype quality information | ADSP QC info | With Multiallelic |
| --- | --- | --- | --- | --- |
| Individual level | CRAMs | - | - | - |
|  | gVCFs | - | - | - |
|  | SV Manta VCFs | - | - | - |
|  | SV Smoove VCFs | - | - | - |
|  | Phenotypes | - | - | - |
| Summary level | Preview VCFs | Full | No | Yes |
|  | Preview Compact VCFs | Partial | No | Yes |
|  | Preview Compact filtered VCFs | Partial | No | Yes |
|  | Fully QC-ed VCFs | Full | Yes | No |
|  | Fully QC-ed Compact filtered VCFs | Partial | Yes | No |
|  | Fully QC-ed GDS | Partial | Yes | No |
|  | Annotation | - | - | - |
|  | LD reference panel | - | - | - |

### Browser of variants and annotations of diversified samples

To allow users to explore the ADSP R4 genotypes without downloading the data, we provide users with two different visualization options. First, VariXam (https://varixam.niagads.org/) allows users to check the alleles and QC quality of any genetic variants in any callsets (this release R4 and earlier ADSP releases) (**Figure 6A**). Users can query by the SNV/gene (IDs or genomic coordinates) level or view all variants in a particular genomic region **(Methods – VariXam)**.

**Figure 6.**
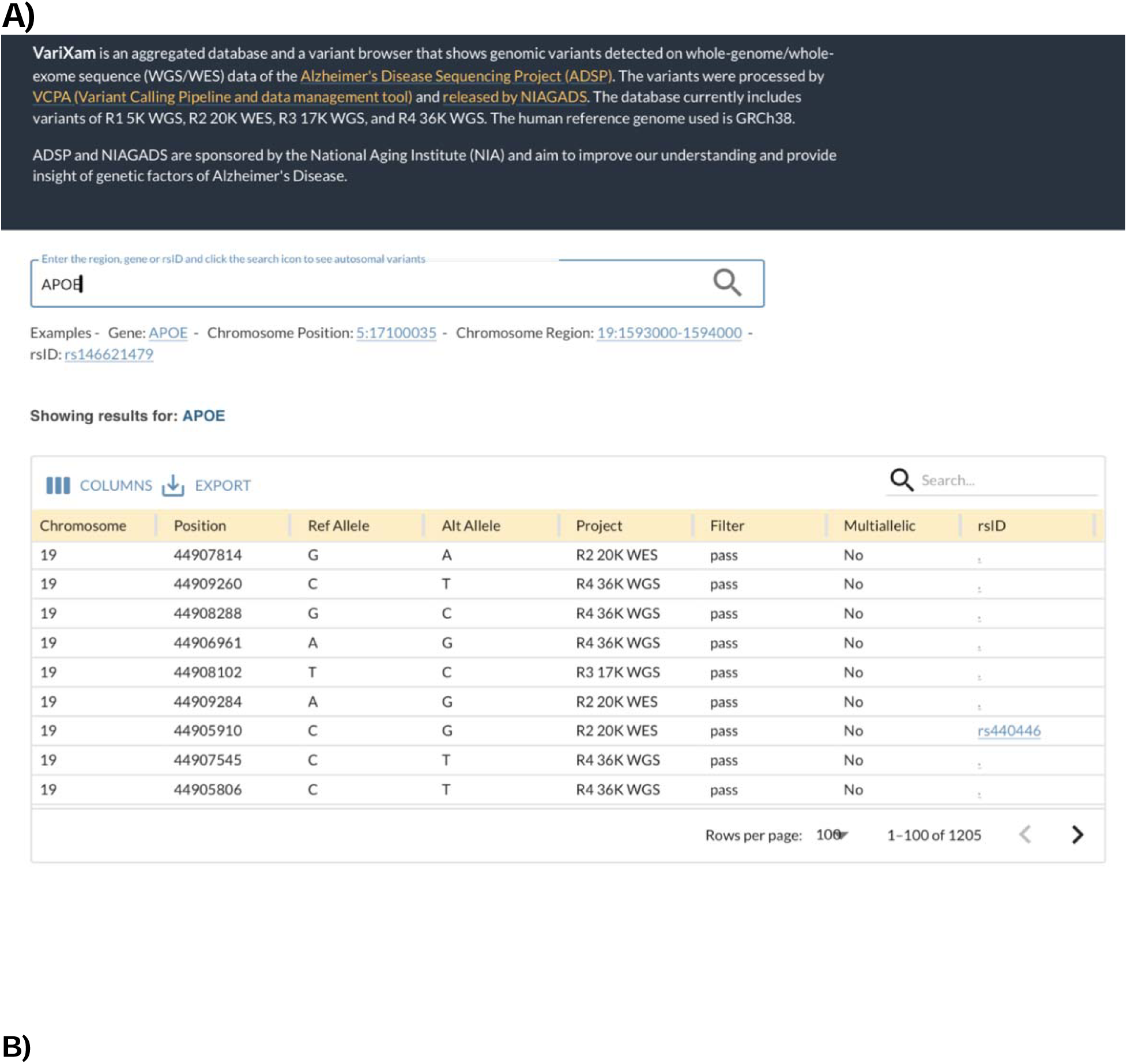
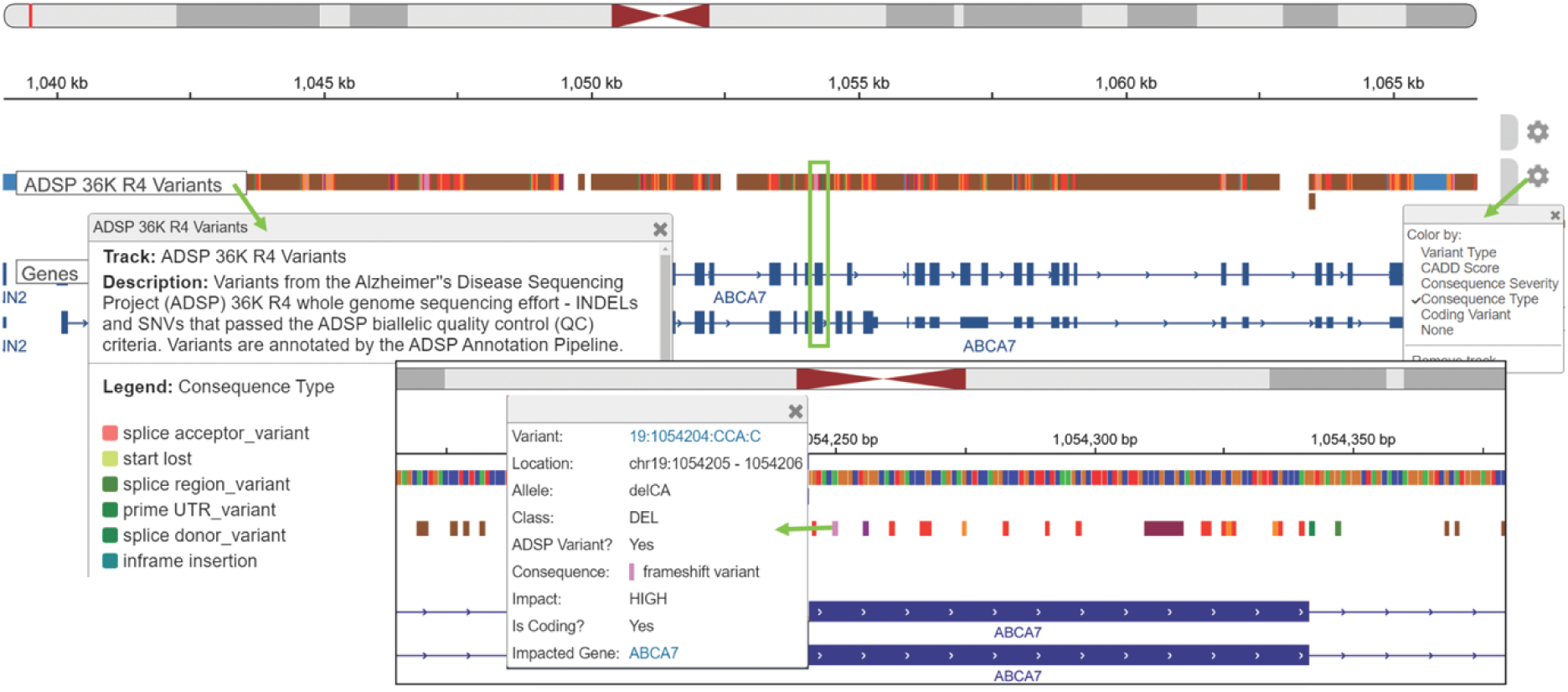
Browser of variants and annotations of diversified samples. **A)** VariXam interface. A variant browser displaying all genomic variants identified in the ADSP whole genome and exome data across releases. The figure below shows the search results of APOE. Accessible at: https://varixam.niagads.org/. **B)** The R4 Variants can be visually inspected as a track on the NIAGADS Genome Browser. The track displays annotated short INDELS and SNVs that passed the biallelic QC criteria. Track annotations include the most severe variant consequences and consequence impacts predicted by the ADSP annotation pipeline and mappings to dbSNP refSNP identifiers. The track settings menu can be used to recolor the variants based on various annotations; the legend (made available by clicking on the track name) will update accordingly. Users can zoom into regions of interest (here, the green rectangle highlights the region displayed in the close-up inset) to view sequence information and click on individual variants for a brief summary of the annotations. Full annotation results can be browsed by following the link to the GenomicsDB record for the variant.

Second, the NIAGADS Alzheimer’s GenomicsDB [27] (https://www.niagads.org/genomics) allows users to explore the variants and annotations in a broader genomic context (**Methods - NIAGADS Alzheimer’s GenomicsDB)**. It provides detailed reports of genetic associations from NIAGADS hosted GWAS summary statistic datasets in the context of genes and annotated variant records. ADSP R4 variants are flagged, and can be filtered by the most severe consequence predicted by the ADSP annotation pipeline [22]. Besides, variant reports also include the full ADSP annotation results (all predicted consequences, CADD [28] deleteriousness, loss of function [29], and FAVOR annotations [29]), allele frequencies (including, gnomAD [20], 1000Genomes [30]), and links out to related web-resources. The GenomicsDB genome browser provides an ADSP R4 variant track that can be recolored based on annotations (e.g., coding vs non-coding variants, consequence type). This track can be compared against the summary statistic data or other tracks in GenomicsDB (**Figure 6B**).

## Discussion

The R4 WGS dataset is the largest and most diverse whole genome data collection for Alzheimer’s disease to date. Numerous studies utilizing the ADSP WGS data in a smaller scale (previous R3 release, 46% of the current sample size, less diversified) have led to multiple findings in AD genetics [31–48], including but not limited to the discovery of 1) new AD genes *PLEC*, *UTRN*, *TP53*, and *POLD1* [33] using a novel approach, GeneEMBED, designed for studying gene interactions; *DLG2* and *DTNB* via rvGWAS on family datasets [31]; 2) rare genetic variants contributing to AD risk [32]; 3) sex-specific loci identified in family-based designs [36]; 4) novel AD risk loci on 13q33.3 via admixture mapping analyses in the Caribbean Hispanic populations [44]; and (5) novel AD associations in Ashkenazi Jews with variants that are exceedingly rare or absent in other European ancestry populations [49]. The ADSP WGS data has also enable the further study of APOE in different ancestries [37, 38, 41, 46, 47] and identification of associations of AD risk in multiple population groups with human viruses detected among unmapped reads in the WGS data [50]. Finally, these data have led to new insights in Early-onset Alzheimer’s (EOAD) [39], enabled the detection of nuclear and mitochondrial copy number variations and structural variants and their association with LOAD [34, 40, 51], as well as the generation of ancestry-specific polygenic risk score in the Amish population [35] and the development of a pipeline for calling mitochondrial sequence variants and haplogroups [52].

We faced several challenges in this project. First, samples are recruited from independent cohorts/projects of various study designs and sequencing experiments are funded during the past 10 years, and challenges rise for GCAD to process and cumulatively integrate all genomic data. Unlike AllofUS [53]UKBiobank [54] the ADSP program has to address the issue of heterogeneity in recruitment criteria, phenotype data collection protocols, and sequencing platform and configuration changes as sequencing technology and analysis best practices continue to evolve. Subtle batch effects may persist in our datasets even after we process all sequence data and perform thorough quality checks during the data harmonization process at sample, variant and phenotype levels.

Second, compared to previous releases (R1 and R3 for genomes), this ADSP R4 dataset more than doubled the sample size (35,014 vs 16,285) with the biggest growth in the HL group (**Supplementary Figure 3**), yet the Asian population is still significantly underrepresented. These gaps must be addressed if we are to fully understand the distribution and effect of human genetic variation in AD. Power analyses show we still need 18,500 cases and 18,500 controls per ancestry group to gain enough power for detecting variants with MAF of 0.005.

In conclusion, we have shown that the ADSP R4 resource offers researchers in the neurodegenerative field multiple tools to explore and analyze the genetic variations associated with these diseases. We anticipate that this data will significantly influence nearly all ongoing studies on common and rare variants in Alzheimer’s disease (AD), with an even greater impact as additional samples from diverse populations become available. Moving forward, we plan to provide annotations for the identified structural variants (SVs) further empowering researchers who depend on such resources to conduct fine-mapping and other post-genome-wide genetic analyses.

The ADSP program needs to share data in a manner that supports the privacy and consent preferences of participants. Members of the scientific community can access most ADSP resources (individual level sequence files, phenotype data, VCFs) through the NIAGADS DSS platform (dbGAP-like). Some companion data (e.g. annotation LD reference panel) are open access, while VariXam variant server and NIAGADS genomicsDB provide a preview of the data without any restrictions.

The ADSP R4 dataset of over 347 million variants is available to qualified investigators at https://dss.niagads.org/datasets/ng00067/.

## Methods

### Sequence existing ancestrally diverse cohorts via the ADSP Follow-Up Study (FUS)

The ADSP-FUS is a NIA initiative focused on identifying genetic risk and protective variants for Alzheimer Disease (AD) by expanding the ADSP cohorts beyond primarily participants with non-Hispanic Whites of European Ancestry (NHW). Given the lack of ancestral diversity in the ADSP, the ADSP-FUS was designed to sequence existing ancestrally diverse and unique cohorts. The current phase for ADSP-FUS, ADSP-FUS 2.0: The Diverse Population Initiative, focuses on Hispanic/Latino (HL), non-Hispanic Black with African Ancestry (NHB-AA), and Asian populations (e.g., the Asian cohort for Alzheimer’s disease). The ADSP-FUS initiatives intend to sequence over 100,000 participants from diverse ancestries. See **Supplementary method** on cohort descriptions. Workflows within the FUS infrastructure (**Supplementary Figure 1**) include biospecimens being processed and DNA prepared and allocated for WGS at USUHS and HIHG. All raw sequence data is transferred to the GCAD for processing and harmonization following quality control (QC) analysis at the University of Pennsylvania and University of Miami, resulting in analysis-ready genotype and sequence data. All clinical, genotype and sequence data are housed at the NIAGADS, which stores, manages, and distributes ASDP-FUS data to AD researchers.

### Dataflow and Sample processing protocol on SNVs and indels

The data flow from receiving data to data sharing is shown in **Supplementary Figure 4**. NIAGADS receives genomes in FASTQ, BAM, or CRAM formats from national or international investigators through secure FTP or S3 bucket. NIAGADS also received the ID information to generate a unique ADSP ID, companion array data for checking concordance, ADSP minimal phenotypes, Institutional Certification forms, and cohort and study information for these genomes. Once the unique ADSP IDs are received and generated by NIAGADS, GCAD can begin production on the genomes.

All genomes are processed using the VCPA pipeline [17] (https://bitbucket.org/NIAGADS/vcpa-pipeline/src/master/), a pipeline optimized for the large-scale production of WGS in an Amazon cloud environment and includes all steps from aligning raw sequence reads to variant calling using GATK best practices. Depending on the file type received, the first few steps of the pipeline are slightly different:

1. CRAM as input: decompression to BAMs is required before running the pipeline as if the input is in BAM format
2. BAM as input: roll back to uBAM (i.e. FASTQ) format before running the pipeline as if the input is in FASTQ format
3. FASTQ as input: no special steps

Using VCPA, FASTQs were first mapped/remapped to hg38 reference genome using BWA-mem (v0.7.15). Duplicated reads were then marked using BamUtil (v1.0.13). Base-recalibration and indel-realignment were done by GATK (v4.1.1) and a CRAM that contains all above information was stored. Genotype calling was then done by GATK4.1.1, resulting in one gVCF across all chromosomes (1-22, X, Y, and M).

Joint genotyping SNVs and indels – we followed the steps in GATK best practices (version GATK4.1.1) to create a joint genotyped called VCFs using gVCFs generated in step 2. VQSR model was performed all chromosomes (1-22, X, Y, and M). This is the “Preview VCF”.

### GRAF-POP

GRAF-pop offers a novel method for global ancestry inference [19], distinguishing itself from traditional approaches that require genotypes from reference populations or study participants to determine an individual’s ancestry. Instead, GRAF-pop precomputes allele frequencies for selected SNPs across reference populations and infers ancestry by directly comparing the test genotypes to these predefined frequencies, eliminating the need for other individual genotypes.

### Population substructure

We included bi-allelic variants that have 1) passed ADSP QC, i.e. with VFLAG=0), 2) Allelic Read Ratio >0.3 or <0.7, 3) MAF>0.02, 4) call rate >99.5% and with hardy-Weinberger test P value larger than 0.0005. Then we applied LD pruning with R cutoff at 0.05, window size of 500K bp. Across all chromosomes, 146,964 pruned variants remained for principal components (PCs)/genetic relationship matrix (GRM) calculations. We used R package “GENESIS” (2.20.1) [55] along with GWASTools(1.36.0) [56] and SNPRelate(1.24.0) [26, 57] to calculate the PCs and GRM. 5513 out of 36361 subjects were grouped as related at default threshold. Note, the threshold value on kinobj used for declaring each pair of participants as related or unrelated. The default value is 2^(−11/2) ∼ 0.022, corresponding to 4th degree relatives.

gnomAD 4.0 genome sites data provided allele frequency at ancestry level, which can be used for ancestry inference. There are 10 ancestries in the data: African/African-American (afr), Amish (ami), Latino (amr), Ashkenazi Jewish (asj), East Asian (eas), Finnish (fin), Middle Eastern (mid), Non-Finnish European (nfe), Remaining, South Asian (sas). 145,278 variants were matched between ADSP pruned file and gnomAD4.0 and passed to Software SCOPE (https://github.com/sriramlab/SCOPE.git) to perform the ancestry proportion analysis. Subject was assigned to ancestry group in which it has the highest ancestry proportion value.

### ADSP Sample level QC protocol

Four checks were performed to identify potentially low-quality samples for exclusion from the VCFs: 1) SNV concordance check with available GWAS genotypes; 2) sex check for possible sample swaps or misreporting; 3) contamination check for possible sample swaps; and 4) relatedness check to confirm known relationships, identify unknown genetically identical duplicates, and assess potential cryptic relatedness.

1. SNV concordance check - this was done by comparing genotypes taken from existing GWAS array data and genotypes from the preview pVCFs on all overlapping genotypes. The goal of this analysis is to ensure that samples and IDs match throughout the data management and calling processes. First, GWAS datasets were converted into VCF format. Then GATK was used to compare the pVCF and GWAS file genotypes using the following parameters: `Java –jar GenomeAnalysisTK.jar –T GenotypeConcordance –R hg38.fa -eval [PVCF] -comp [GWAS_VCF] -o [OUTFILE]` where [GWAS_VCF] is the VCF file converted from GWAS, [PVCF] is the Preview pVCF, and [OUTFILE] is the user-selected output filename. Of all samples kept, they all have a concordance of > 0.91.
2. Sex checks were performed using BCFtools [58], VCFtools [59] and PLINK [60] with the following steps:

a. Use bcftools to convert chromosome X pvcf into plink format
b. Filter out chromosome X pseudo-autosomal region (PAR)
c. Filter out SNVs with MAF<0.05, run ‘impute-sex’
d. Run sex-check using PLINK for comparison Comparing the results of ‘impute-sex’ in BCFtools and ‘’--sex-check” in PLINK without filtering on minor allele frequencies (MAFs) or without excluding variants in the pseudoautosomal region (PAR), the findings of the two approaches are the same. There are 1,918 samples with F-statistic values between 0.2 and 0.8, but only 392 between 0.3 and 0.7. A total of 74 samples were identified to be the incorrect sex. The submitting centers agreed that these subjects needed to be dropped.
3. Sample-specific contamination checking was performed by using VerifyBamID [61] to calculate the concordance estimate between the GWAS genotype data and the BAM file. This approach provides information that can be interpreted to identify potential sample contamination or swapping using the GWAS-BAM contamination estimate. The ‘FREEMIX’ modeling approach was used in this analysis. We used suggested thresholds for contamination taken from the VerifyBamID website: a sample is potential contaminated if the FREEMIX value is >0.05. The command line to generate FREEMIX contamination estimates value is as follows: `verifyBamID --vcf [INPUT.VCF] --bam [INPUT.BAM] -- out [OUTPUT.PREFIX] --verbose --ignoreRG` where [INPUT.VCF] is the VCF file converted from GWAS, [INPUT.BAM] is the BAM file generated using VCPA1.1, and [OUTPUT.PREFIX] is the user-selected prefix to be appended to output files. As implemented, DNA sample contamination is indicated if the FREEMIX value is >0.05. Across all samples, all FREEMIX values obtained are <0.05.
4. Relatedness checks were performed using PLINK as described:

a. Convert *vcf.gz files into PLINK binary format - Run PLINK “--recode” and “--make-bed” commands to convert ‘*.vcf.gz’ files into ‘*.bed’, ‘*.bim’, and ‘*.fam’ files
b. Combine 22 chromosome-specific PLINK filesets into one genome-wide set and organize

- Clean and rename empty name “.” snvs to “chr_pos” format to combine
- Run PLINK “--merge-list” and “--make-bed” commands to combine 22 binary files into one
- Run PLINK “—extract” command to extract only overlapping ∼21k SNVs
c. Run PLINK to generate pairwise sample IBD values

- Run PLINK “--genome” command on combined file with ∼21k SNVs to get pairwise IBD values
- Using these IBD values, identify related pairs with IBD PI_HAT > 0.4 The PI_HAT measurement [Proportion IBD, i.e. P(IBD=2) + 0.5*P(IBD=1)] was reported for each pair of samples. All pairs with PI_HAT > 0.4 were evaluated for known relatedness.

There was a total of 1,205 genetically unique participants identified with multiple samples (including 5 pairs of twins) across the R4 dataset. This included 6 known replicates (sequenced a total of 67 times as technical replicates), and 1,199 unintentional replicates (1,173 pairs and 26 triplicates). After removing the ADSP recommended duplicate samples, there were a total of 35,023 genetically unique samples. NOTE: there are 24 subjects from the 36k listed in both the family-based and case/control phenotype files. These samples were either sequenced 1) as part of a case/control study but were also part of a family so their phenotypes are provided in both files, or 2) in both a case/control and family-based study. ADSP recommends using samples from family-based sets.

After performing sex check, GWAS concordance and contamination checks for each sample, together with the metrics we collected per sample in VCPA, we designed a list of criteria to drop or fail samples based on this sample level QC process (**Supplementary Figure 5**) –. A sample is dropped if a) its average genome coverage is lower than 20; or b) multiple of its non-related sequencing metrics are of bad quality; or c) it fails contamination check, concordance check, or sex check; or 4) it is an unexpected duplicate. Alternatively, a sample is flagged if fails in less than 3 sequencing metrics.

### ADSP Variant QC protocol

Different filtering and quality-checking strategies were applied at each level (genotype-, and variant-) [62]. The QC protocol was applied on the bi-allelic autosomal VCFs. All QC flags were applied uniformly across all samples, regardless of cohort or sequencing information. Variants of low quality have been flagged but these variants have not been excluded and filtered out of the datasets.

1. Genotype-level QC was applied to individual genotypes. Each genotype was evaluated and set to missing (“./.”) if either or both read depth (“DP”) was less than 10 (DP<10) or genotype quality (“GQ”) score was less than 20 (GQ<20). All these censored genotypes were excluded from subsequent QC steps, except for estimation of variant-level averaged depth (“AverageReadDepth”) in variant-level QC.
2. Variant-level QC was applied to all variants. Flags were applied in the following order: a) Variants in GATK low sequence quality tranches [variants without a FILTER value of “PASS” that are above the 99.8% VQSR Tranche]; b) Monomorphic variants were flagged; c) Variants with high missing rate were flagged; d) Variants with high read depth were flagged.
3. Variants with excessive heterozygosity or departure from Hardy-Weinberg equilibrium (HWE) were evaluated within race/ethnic subgroup, however given the complexity around race/ethnic subgroups, these were not flagged though the measures have been made available and can be implemented as user-defined filters if desired. Similarly, (“ABhet”) was computed among uncensored heterozygotes at each variant and provided in the files but not applied in any filtering criteria.

### Comparison with gnomAD

The gnomAD resource (version 4) were download from https://gnomad.broadinstitute.org/downloads#v4. This contains bi-allelic variants from multi-ethnic samples. To compare with the ADSP R4 data, we first extracted the bi-allelic variants from the VCF. We then broke the VCFs down into samples of four ancestry groups – NHW, HL, Asians and HHB-AA. Monomorphic variants were excluded from each of the VCFs. We then extracted variants from both the gnomAD and ADSP R4 data by MAF thresholds: <=0.1%, <=0.5%, <=1% and <=5%, and compared them at the site level. When compared across ancestry groups, only sites that are observed in all ancestries were used for analyses.

### Variant annotation protocol

All R4 bi-allelic variants have been annotated using the official ADSP annotation pipeline. First, the QCed VCFs were processed using VEP103 [63] (with the --everything flag). Then the JSON-formatted VEP output was processed so that variants affecting multiple transcripts of the same gene were collapsed generate a ‘most damaging’ consequence for each affected gene. This process uses the ranking table specified in the file ‘ranking_table.txt’ to identify the ‘most damaging’ consequence and to assign an impact score, using a custom annotation ranking process which down-weights consequences for non-sense mediated decay transcripts and non-coding transcripts. Next the QCed VCFs were also processed by SnpEff v5.1d (build 2022-04-19) [23]. Variants are matched by chromosome, position, reference allele, and alternate allele to CADD v1.6 scores [28]. Lastly, short indels not defined in CADD reference files were processed by CADD and integrated into the dataset. This resource is available at NIAGADS open access https://dss.niagads.org/open-access-data-portal/.

### LOF analyses

The functional impact of variants was assessed using SnpEff [23]. Variants were annotated as ‘MODERATE’ and ‘HIGH’ when they were protein-altering, while variants with ‘LOW’ and ‘MODIFIER’ effects were considered non-protein altering. All variants categorized as ‘HIGH’ were expected to be disruptive or cause loss of function (LoF) in the protein. Variants with a ‘MODERATE’ effect were expected to be missense and splice region variants. Variants with ‘MODIFIER’ and ‘LOW’ effects were in non-coding regions or were non-disruptive to protein functions. We focused on the LoF variants categorized as ‘HIGH’ in the ADSP cohort.

### FAVOR annotation protocol

We downloaded the FAVOR [29] database annotations from https://docs.genohub.org/data in July 2023 and used that to annotate all the R4 bi-allelic variants. Then annotation only VCF files were converted to GDS format using the SeqArray package [26], containing 156 columns. This resource is available at NIAGADS open access https://dss.niagads.org/open-access-data-portal/.

### Structural variant calling protocol

The GCAD and ADSP SV workgroup together designs the production pipeline which includes Manta [64] (v1.6.0) and Smoove (ref, v0.2.6) (https://github.com/brentp/smoove) for calling deletions and insertions. Individual Manta and Smoove callsets were first merged for each sample and merged together with all samples by SVIMMER (v0.1) (https://github.com/DecodeGenetics/svimmer). Then, GraphTyper (v2.7) [65] was applied on the merged VCF for SV joint genotyping. Note that the only filter applied is SV size >10 Mbp; other than that, there is no advance filter. The code used to generate these SV calls is available at: https://github.com/Illumina/manta, https://github.com/brentp/smoove).

### LD reference panel

We inferred LD separately for each of the four major ancestral groups (NHW, His, AA, and Asian). All participants for both cases and controls were included but PSP and CBD samples are not included, resulting in 32,236 participants total). We calculated LD for all pairs of variants with minor allele count (MAC) ≥ 5 and within 5 Mbps of each other using emerald [25] using the following parameters, --mac > 5, --threshold 0.2, and --window 5000000. Each segment was analyzed by 5Mb window with a 3Mb overlapping, then we concatenated all segments and removed duplicate records. Only variant pairs with R > 0.2 were then retained. For each variant pair, we reported variant genomic positions, reference and non-reference alleles, their R and R correlation, and D and D’ statistics. This resource is available at NIAGADS open access https://dss.niagads.org/open-access-data-portal/.

### ADSP Phenotype Harmonization Consortium (ADSP-PHC)

The ADSP Phenotype Harmonization Consortium (ADSP-PHC) harmonizes all available data from each domain, regardless of sequencing status, to ensure the highest quality harmonization. The harmonized phenotypic data are then subset to participants with available whole-genome sequencing. These data are released per participant via NIAGADS. All harmonized data can be accessed directly from each cohort. All ADSP phenotype data are harmonized by a multi-disciplinary team that includes world experts in neuroimaging, neuropsychology, fluid biomarkers, neuropathology, and vascular contributions to ADRD. Data processing and domain-specific harmonization protocols are available in Supplementary Methods - ADSP Phenotype Harmonization Consortium Protocol. These files are available in https://dss.niagads.org/.

### VariXam

This is an aggregated database and a variant browser that shows genomic variants detected on WGS/WES data of the ADSP. The database currently includes variants of all the R4 36K WGS and allows users to search for genes or variants of interests. The human reference genome used is GRCh38. It is available here: https://varixam.niagads.org/.

### NIAGADS Alzheimer’s GenomicsDB

This is an interactive knowledgebase for AD genetics [27]. The resource provides unrestricted access to GWAS summary statistics datasets, variant annotations, and meta-analysis results deposited at the NIAGADS. The platform allows users to search for genes or variants of interests, and interactively mine or visually inspect datasets and annotated ADSP variant tracks on a genome browser. The GenomicsDB can be accessed at https://www.niagads.org/genomics.

### NIAGADS DSS

The NIAGADS Data Sharing Service (DSS) was developed to facilitate the deposition and sharing of whole-genome and whole-exome sequencing data from ADSP and other NIA funded ADRD studies to the research community at large. In keeping with the NIH Genomic Data Sharing (GDS) Policy, all genomic data are classified as controlled access as outlined in the Institutional Certification forms provided by the submitting institutions. Principal investigators can request DSS distributed data through the Data Access Request Management (DARM) system by logging in using their eRA Commons ID. Once an application is approved by the NIH-formed NIAGADS ADRD Data Access Committee (NADAC) and Data Use Committee (DUC), the data can be accessed through the Data Portal and downloaded directly or through Amazon EC2. DSS can be found at https://dss.niagads.org/.

### Released genotyping files

Due to the sheer sizes of pVCFs, all pVCFs are split by chromosomes. We provide three versions of pVCFs for users to choose from: (1) “Preview pVCF”; (2) “Compact pVCF”: only reserved GT (genotype) of each sample for each variant; (3) “Compact filtered pVCF”: a compact version with replacing low-quality genotypes to missing (./.). Each set of pVCF files are divided by chromosome, then split into bi-allelic and multi-allelic variant files. Besides, we also created the fully QCed VCF (output from the “ADSP Variant QC protocol”) in Genomic Data Structure (GDS) format to facilitate analysts using R for downstream association analyses. These files are available in https://dss.niagads.org/.

## Data Availability

All data described are available at https://dss.niagads.org/. VariXam is accessible at https://varixam.niagads.org/. GenomicsDB is accessible at https://www.niagads.org/genomics/app.

## Authors’ contribution

YYL, W-PL, LAF, RPM, MAP-V, JLH, EM, ACN, XZ, BNV, BWK, WSB, GDS, and L-SW conceptualized the study. CD, LAF, RPM, MAP-V, JLH, EM, AT, MC, TJH, GDS, and L-SW secures the funding. YYL, ABK, HN, HW, ZK, LC, PRM, CD, THJ, and BWK performed data management and coordination of the project. YYL, OV, PG, LQ, YZ, P-LC, HN, LA, NS, TI, and NRW generated the resources. YYL, W-PL, PPK, TIG, CZ, HW, JJ, and WSB performed the analyses. YYL, W-PK, PPK, SLT, TIG, CAN, XZ, BNV, TJH, HW, JJ, BWK, and WSB drafted the manuscript.

## Acknowledgements

Data for this study were prepared, archived, and distributed by the National Institute on Aging Alzheimer’s Disease Data Storage Site (NIAGADS) at the University of Pennsylvania (U24-AG041689), funded by the National Institute on Aging.

## Alzheimer’s Disease Sequencing Project (sa000001) data

The Alzheimer’s Disease Sequencing Project (ADSP) is comprised of two Alzheimer’s Disease (AD) genetics consortia and three National Human Genome Research Institute (NHGRI) funded Large Scale Sequencing and Analysis Centers (LSAC). The two AD genetics consortia are the Alzheimer’s Disease Genetics Consortium (ADGC) funded by NIA (U01 AG032984), and the Cohorts for Heart and Aging Research in Genomic Epidemiology (CHARGE) funded by NIA (R01 AG033193), the National Heart, Lung, and Blood Institute (NHLBI), other National Institute of Health (NIH) institutes and other foreign governmental and non-governmental organizations. The Discovery Phase analysis of sequence data is supported through UF1AG047133 (to Drs. Schellenberg, Farrer, Pericak-Vance, Mayeux, and Haines); U01AG049505 to Dr. Seshadri; U01AG049506 to Dr. Boerwinkle; U01AG049507 to Dr. Wijsman; and U01AG049508 to Dr. Goate and the Discovery Extension Phase analysis is supported through U01AG052411 to Dr. Goate, U01AG052410 to Dr. Pericak-Vance and U01 AG052409 to Drs. Seshadri and Fornage.

Sequencing for the Follow Up Study (FUS) is supported through U01AG057659 (to Drs. PericakVance, Mayeux, and Vardarajan) and U01AG062943 (to Drs. Pericak-Vance and Mayeux). Data generation and harmonization in the Follow-up Phase is supported by U54AG052427 (to Drs. Schellenberg and Wang). The FUS Phase analysis of sequence data is supported through U01AG058589 (to Drs. Destefano, Boerwinkle, De Jager, Fornage, Seshadri, and Wijsman), U01AG058654 (to Drs. Haines, Bush, Farrer, Martin, and Pericak-Vance), U01AG058635 (to Dr. Goate), RF1AG058066 (to Drs. Haines, Pericak-Vance, and Scott), RF1AG057519 (to Drs. Farrer and Jun), R01AG048927 (to Dr. Farrer), and RF1AG054074 (to Drs. Pericak-Vance and Beecham).

The ADGC cohorts include: Adult Changes in Thought (ACT) (U01 AG006781, U19 AG066567), the Alzheimer’s Disease Research Centers (ADRC) (P30 AG062429, P30 AG066468, P30 AG062421, P30 AG066509, P30 AG066514, P30 AG066530, P30 AG066507, P30 AG066444, P30 AG066518, P30 AG066512, P30 AG066462, P30 AG072979, P30 AG072972, P30 AG072976, P30 AG072975, P30 AG072978, P30 AG072977, P30 AG066519, P30 AG062677, P30 AG079280, P30 AG062422, P30 AG066511, P30 AG072946, P30 AG062715, P30 AG072973, P30 AG066506, P30 AG066508, P30 AG066515, P30 AG072947, P30 AG072931, P30 AG066546, P20 AG068024, P20 AG068053, P20 AG068077, P20 AG068082, P30 AG072958, P30 AG072959), the Chicago Health and Aging Project (CHAP) (R01 AG11101, RC4 AG039085, K23 AG030944), Indiana Memory and Aging Study (IMAS) (R01 AG019771), Indianapolis Ibadan (R01 AG009956, P30 AG010133), the Memory and Aging Project (MAP) ( R01 AG17917), Mayo Clinic (MAYO) (R01 AG032990, U01 AG046139, R01 NS080820, RF1 AG051504, P50 AG016574), Mayo Parkinson’s Disease controls (NS039764, NS071674, 5RC2HG005605), University of Miami (R01 AG027944, R01 AG028786, R01 AG019085, IIRG09133827, A2011048), the Multi-Institutional Research in Alzheimer’s Genetic Epidemiology Study (MIRAGE) (R01 AG09029, R01 AG025259), the National Centralized Repository for Alzheimer’s Disease and Related Dementias (NCRAD) (U24 AG021886), the National Institute on Aging Late Onset Alzheimer’s Disease Family Study (NIA-LOAD) (U24 AG056270), the Religious Orders Study (ROS) (P30 AG10161, R01 AG15819), the Texas Alzheimer’s Research and Care Consortium (TARCC) (funded by the Darrell K Royal Texas Alzheimer’s Initiative), Vanderbilt University/Case Western Reserve University (VAN/CWRU) (R01 AG019757, R01 AG021547, R01 AG027944, R01 AG028786, P01 NS026630, and Alzheimer’s Association), the Washington Heights-Inwood Columbia Aging Project (WHICAP) (RF1 AG054023), the University of Washington Families (VA Research Merit Grant, NIA: P50AG005136, R01AG041797, NINDS: R01NS069719), the Columbia University Hispanic Estudio Familiar de Influencia Genetica de Alzheimer (EFIGA) (RF1 AG015473), the University of Toronto (UT) (funded by Wellcome Trust, Medical Research Council, Canadian Institutes of Health Research), and Genetic Differences (GD) (R01 AG007584). The CHARGE cohorts are supported in part by National Heart, Lung, and Blood Institute (NHLBI) infrastructure grant HL105756 (Psaty), RC2HL102419 (Boerwinkle) and the neurology working group is supported by the National Institute on Aging (NIA) R01 grant AG033193.

The CHARGE cohorts participating in the ADSP include the following: Austrian Stroke Prevention Study (ASPS), ASPS-Family study, and the Prospective Dementia Registry-Austria (ASPS/PRODEM-Aus), the Atherosclerosis Risk in Communities (ARIC) Study, the Cardiovascular Health Study (CHS), the Erasmus Rucphen Family Study (ERF), the Framingham Heart Study (FHS), and the Rotterdam Study (RS). ASPS is funded by the Austrian Science Fond (FWF) grant number P20545-P05 and P13180 and the Medical University of Graz. The ASPS-Fam is funded by the Austrian Science Fund (FWF) project I904), the EU Joint Programme – Neurodegenerative Disease Research (JPND) in frame of the BRIDGET project (Austria, Ministry of Science) and the Medical University of Graz and the Steiermärkische Krankenanstalten Gesellschaft. PRODEM-Austria is supported by the Austrian Research Promotion agency (FFG) (Project No. 827462) and by the Austrian National Bank (Anniversary Fund, project 15435. ARIC research is carried out as a collaborative study supported by NHLBI contracts (HHSN268201100005C, HHSN268201100006C, HHSN268201100007C, HHSN268201100008C, HHSN268201100009C, HHSN268201100010C, HHSN268201100011C, and HHSN268201100012C). Neurocognitive data in ARIC is collected by U01 2U01HL096812, 2U01HL096814, 2U01HL096899, 2U01HL096902, 2U01HL096917 from the NIH (NHLBI, NINDS, NIA and NIDCD), and with previous brain MRI examinations funded by R01-HL70825 from the NHLBI. CHS research was supported by contracts HHSN268201200036C, HHSN268200800007C, N01HC55222, N01HC85079, N01HC85080, N01HC85081, N01HC85082, N01HC85083, N01HC85086, and grants U01HL080295 and U01HL130114 from the NHLBI with additional contribution from the National Institute of Neurological Disorders and Stroke (NINDS). Additional support was provided by R01AG023629, R01AG15928, and R01AG20098 from the NIA. FHS research is supported by NHLBI contracts N01-HC-25195 and HHSN268201500001I. This study was also supported by additional grants from the NIA (R01s AG054076, AG049607 and AG033040 and NINDS (R01 NS017950). The ERF study as a part of EUROSPAN (European Special Populations Research Network) was supported by European Commission FP6 STRP grant number 018947 (LSHG-CT-2006-01947) and also received funding from the European Community’s Seventh Framework Programme (FP7/2007-2013)/grant agreement HEALTH-F4-2007-201413 by the European Commission under the programme “Quality of Life and Management of the Living Resources” of 5th Framework Programme (no. QLG2-CT-2002-01254). High-throughput analysis of the ERF data was supported by a joint grant from the Netherlands Organization for Scientific Research and the Russian Foundation for Basic Research (NWO-RFBR 047.017.043). The Rotterdam Study is funded by Erasmus Medical Center and Erasmus University, Rotterdam, the Netherlands Organization for Health Research and Development (ZonMw), the Research Institute for Diseases in the Elderly (RIDE), the Ministry of Education, Culture and Science, the Ministry for Health, Welfare and Sports, the European Commission (DG XII), and the municipality of Rotterdam. Genetic data sets are also supported by the Netherlands Organization of Scientific Research NWO Investments (175.010.2005.011, 911-03-012), the Genetic Laboratory of the Department of Internal Medicine, Erasmus MC, the Research Institute for Diseases in the Elderly (014-93-015; RIDE2), and the Netherlands Genomics Initiative (NGI)/Netherlands Organization for Scientific Research (NWO) Netherlands Consortium for Healthy Aging (NCHA), project 050-060-810. All studies are grateful to their participants, faculty and staff. The content of these manuscripts is solely the responsibility of the authors and does not necessarily represent the official views of the National Institutes of Health or the U.S. Department of Health and Human Services.

The FUS cohorts include: the Alzheimer’s Disease Research Centers (ADRC) (P30 AG062429, P30 AG066468, P30 AG062421, P30 AG066509, P30 AG066514, P30 AG066530, P30 AG066507, P30 AG066444, P30 AG066518, P30 AG066512, P30 AG066462, P30 AG072979, P30 AG072972, P30 AG072976, P30 AG072975, P30 AG072978, P30 AG072977, P30 AG066519, P30 AG062677, P30 AG079280, P30 AG062422, P30 AG066511, P30 AG072946, P30 AG062715, P30 AG072973, P30 AG066506, P30 AG066508, P30 AG066515, P30 AG072947, P30 AG072931, P30 AG066546, P20 AG068024, P20 AG068053, P20 AG068077, P20 AG068082, P30 AG072958, P30 AG072959), Alzheimer’s Disease Neuroimaging Initiative (ADNI) (U19AG024904), Amish Protective Variant Study (RF1AG058066), Cache County Study (R01AG11380, R01AG031272, R01AG21136, RF1AG054052), Case Western Reserve University Brain Bank (CWRUBB) (P50AG008012), Case Western Reserve University Rapid Decline (CWRURD) (RF1AG058267, NU38CK000480), CubanAmerican Alzheimer’s Disease Initiative (CuAADI) (3U01AG052410), Estudio Familiar de Influencia Genetica en Alzheimer (EFIGA) (5R37AG015473, RF1AG015473, R56AG051876), Genetic and Environmental Risk Factors for Alzheimer Disease Among African Americans Study (GenerAAtions) (2R01AG09029, R01AG025259, 2R01AG048927), Gwangju Alzheimer and Related Dementias Study (GARD) (U01AG062602), Hillblom Aging Network (2014-A-004-NET, R01AG032289, R01AG048234), Hussman Institute for Human Genomics Brain Bank (HIHGBB) (R01AG027944, Alzheimer’s Association “Identification of Rare Variants in Alzheimer Disease”), Ibadan Study of Aging (IBADAN) (5R01AG009956), Longevity Genes Project (LGP) and LonGenity (R01AG042188, R01AG044829, R01AG046949, R01AG057909, R01AG061155, P30AG038072), Mexican Health and Aging Study (MHAS) (R01AG018016), Multi-Institutional Research in Alzheimer’s Genetic Epidemiology (MIRAGE) (2R01AG09029, R01AG025259, 2R01AG048927), Northern Manhattan Study (NOMAS) (R01NS29993), Peru Alzheimer’s Disease Initiative (PeADI) (RF1AG054074), Puerto Rican 1066 (PR1066) (Wellcome Trust (GR066133/GR080002), European Research Council (340755)), Puerto Rican Alzheimer Disease Initiative (PRADI) (RF1AG054074), Reasons for Geographic and Racial Differences in Stroke (REGARDS) (U01NS041588), Research in African American Alzheimer Disease Initiative (REAAADI) (U01AG052410), the Religious Orders Study (ROS) (P30 AG10161, P30 AG72975, R01 AG15819, R01 AG42210), the RUSH Memory and Aging Project (MAP) (R01 AG017917, R01 AG42210Stanford Extreme Phenotypes in AD (R01AG060747), University of Miami Brain Endowment Bank (MBB), University of Miami/Case Western/North Carolina A&T African American (UM/CASE/NCAT) (U01AG052410, R01AG028786), and Wisconsin Registry for Alzheimer’s Prevention (WRAP) (R01AG027161 and R01AG054047).

The four LSACs are: the Human Genome Sequencing Center at the Baylor College of Medicine (U54 HG003273), the Broad Institute Genome Center (U54HG003067), The American Genome Center at the Uniformed Services University of the Health Sciences (U01AG057659), and the Washington University Genome Institute (U54HG003079). Genotyping and sequencing for the ADSP FUS is also conducted at John P. Hussman Institute for Human Genomics (HIHG) Center for Genome Technology (CGT).

Biological samples and associated phenotypic data used in primary data analyses were stored at Study Investigators institutions, and at the National Centralized Repository for Alzheimer’s Disease and Related Dementias (NCRAD, U24AG021886) at Indiana University funded by NIA. Associated Phenotypic Data used in primary and secondary data analyses were provided by Study Investigators, the NIA funded Alzheimer’s Disease Centers (ADCs), and the National Alzheimer’s Coordinating Center (NACC, U24AG072122) and the National Institute on Aging Genetics of Alzheimer’s Disease Data Storage Site (NIAGADS, U24AG041689) at the University of Pennsylvania, funded by NIA. Harmonized phenotypes were provided by the ADSP Phenotype Harmonization Consortium (ADSP-PHC), funded by NIA (U24 AG074855, U01 AG068057 and R01 AG059716) and Ultrascale Machine Learning to Empower Discovery in Alzheimer’s Disease Biobanks (AI4AD, U01 AG068057). This research was supported in part by the Intramural Research Program of the National Institutes of health, National Library of Medicine. Contributors to the Genetic Analysis Data included Study Investigators on projects that were individually funded by NIA, and other NIH institutes, and by private U.S. organizations, or foreign governmental or nongovernmental organizations.

The ADSP Phenotype Harmonization Consortium (ADSP-PHC) is funded by NIA (U24 AG074855, U01 AG068057 and R01 AG059716). The harmonized cohorts within the ADSP-PHC include:1the Anti-Amyloid Treatment in Asymptomatic Alzheimer’s study (A4 Study), a secondary prevention trial in preclinical Alzheimer’s disease, aiming to slow cognitive decline associated with brain amyloid accumulation in clinically normal older participants. The A4 Study is funded by a public-private-philanthropic partnership, including funding from the National Institutes of Health-National Institute on Aging, Eli Lilly and Company, Alzheimer’s Association, Accelerating Medicines Partnership, GHR Foundation, an anonymous foundation and additional private donors, with in-kind support from Avid and Cogstate. The companion observational Longitudinal Evaluation of Amyloid Risk and Neurodegeneration (LEARN) Study is funded by the Alzheimer’s Association and GHR Foundation. The A4 and LEARN Studies are led by Dr. Reisa Sperling at Brigham and Women’s Hospital, Harvard Medical School and Dr. Paul Aisen at the Alzheimer’s Therapeutic Research Institute (ATRI), University of Southern California. The A4 and LEARN Studies are coordinated by ATRI at the University of Southern California, and the data are made available through the Laboratory for Neuro Imaging at the University of Southern California. The participants screening for the A4 Study provided permission to share their de-identified data in order to advance the quest to find a successful treatment for Alzheimer’s disease. We would like to acknowledge the dedication of all the participants, the site personnel, and all of the partnership team members who continue to make the A4 and LEARN Studies possible. The complete A4 Study Team list is available on: a4study.org<u>/a4-study-team.</u>; the Adult Changes in Thought study (ACT), U01 AG006781, U19 AG066567; Alzheimer’s Disease Neuroimaging Initiative (ADNI): Data collection and sharing for this project was funded by the Alzheimer’s Disease Neuroimaging Initiative (ADNI) (National Institutes of Health Grant U01 AG024904) and DOD ADNI (Department of Defense award number W81XWH-12-2-0012). ADNI is funded by the National Institute on Aging, the National Institute of Biomedical Imaging and Bioengineering, and through generous contributions from the following: AbbVie, Alzheimer’s Association; Alzheimer’s Drug Discovery Foundation; Araclon Biotech; BioClinica, Inc.; Biogen; Bristol-Myers Squibb Company; CereSpir, Inc.; Cogstate; Eisai Inc.; Elan Pharmaceuticals, Inc.; Eli Lilly and Company; EuroImmun; F. Hoffmann-La Roche Ltd and its affiliated company Genentech, Inc.; Fujirebio; GE Healthcare; IXICO Ltd.;Janssen Alzheimer Immunotherapy Research & Development, LLC.; Johnson & Johnson Pharmaceutical Research & Development LLC.; Lumosity; Lundbeck; Merck & Co., Inc.;Meso Scale Diagnostics, LLC.; NeuroRx Research; Neurotrack Technologies; Novartis Pharmaceuticals Corporation; Pfizer Inc.; Piramal Imaging; Servier; Takeda Pharmaceutical Company; and Transition Therapeutics. The Canadian Institutes of Health Research is providing funds to support ADNI clinical sites in Canada. Private sector contributions are facilitated by the Foundation for the National Institutes of Health (www.fnih.org). The grantee organization is the Northern California Institute for Research and

Education, and the study is coordinated by the Alzheimer’s Therapeutic Research Institute at the University of Southern California. ADNI data are disseminated by the Laboratory for Neuro Imaging at the University of Southern California; Estudio Familiar de Influencia Genetica en Alzheimer (EFIGA): 5R37AG015473, RF1AG015473, R56AG051876; Memory & Aging Project at Knight Alzheimer’s Disease Research Center (MAP at Knight ADRC): The Memory and Aging Project at the Knight-ADRC (Knight-ADRC). This work was supported by the National Institutes of Health (NIH) grants R01AG064614, R01AG044546, RF1AG053303, RF1AG058501, U01AG058922 and R01AG064877 to Carlos Cruchaga. The recruitment and clinical characterization of research participants at Washington University was supported by NIH grants P30AG066444, P01AG03991, and P01AG026276. Data collection and sharing for this project was supported by NIH grants RF1AG054080, P30AG066462, R01AG064614 and U01AG052410. We thank the contributors who collected samples used in this study, as well as patients and their families, whose help and participation made this work possible. This work was supported by access to equipment made possible by the Hope Center for Neurological Disorders, the Neurogenomics and Informatics Center (NGI: https://neurogenomics.wustl.edu/) and the Departments of Neurology and Psychiatry at Washington University School of Medicine; National Alzheimer’s Coordinating Center (NACC): The NACC database is funded by NIA/NIH Grant U24 AG072122. NACC data are contributed by the NIA-funded ADRCs: P30 AG062429 (PI James Brewer, MD, PhD), P30 AG066468 (PI Oscar Lopez, MD), P30 AG062421 (PI Bradley Hyman, MD, PhD), P30 AG066509 (PI Thomas Grabowski, MD), P30 AG066514 (PI Mary Sano, PhD), P30 AG066530 (PI Helena Chui, MD), P30 AG066507 (PI Marilyn Albert, PhD), P30 AG066444 (PI John Morris, MD), P30 AG066518 (PI Jeffrey Kaye, MD), P30 AG066512 (PI Thomas Wisniewski, MD), P30 AG066462 (PI Scott Small, MD), P30 AG072979 (PI David Wolk, MD), P30 AG072972 (PI Charles DeCarli, MD), P30 AG072976 (PI Andrew Saykin, PsyD), P30 AG072975 (PI David Bennett, MD), P30 AG072978 (PI Neil Kowall, MD), P30 AG072977 (PI Robert Vassar, PhD), P30 AG066519 (PI Frank LaFerla, PhD), P30 AG062677 (PI Ronald Petersen, MD, PhD), P30 AG079280 (PI Eric Reiman, MD), P30 AG062422 (PI Gil Rabinovici, MD), P30 AG066511 (PI Allan Levey, MD, PhD), P30 AG072946 (PI Linda Van Eldik, PhD), P30 AG062715 (PI Sanjay Asthana, MD, FRCP), P30 AG072973 (PI Russell Swerdlow, MD), P30 AG066506 (PI Todd Golde, MD, PhD), P30 AG066508 (PI Stephen Strittmatter, MD, PhD), P30 AG066515 (PI Victor Henderson, MD, MS), P30 AG072947 (PI Suzanne Craft, PhD), P30 AG072931 (PI Henry Paulson, MD, PhD), P30 AG066546 (PI Sudha Seshadri, MD), P20 AG068024 (PI Erik Roberson, MD, PhD), P20 AG068053 (PI Justin Miller, PhD), P20 AG068077 (PI Gary Rosenberg, MD), P20 AG068082 (PI Angela Jefferson, PhD), P30 AG072958 (PI Heather Whitson, MD), P30 AG072959 (PI James Leverenz, MD); National Institute on Aging Alzheimer’s Disease Family Based Study (NIA-AD FBS): U24 AG056270; Religious Orders Study (ROS): P30AG10161,R01AG15819, R01AG42210; Memory and Aging Project (MAP - Rush): R01AG017917, R01AG42210; Minority Aging Research Study (MARS): R01AG22018, R01AG42210; Washington Heights/Inwood Columbia Aging Project (WHICAP): RF1 AG054023;and Wisconsin Registry for Alzheimer’s Prevention (WRAP): R01AG027161 and R01AG054047. Additional acknowledgments include the National Institute on Aging Genetics of Alzheimer’s Disease Data Storage Site (NIAGADS, U24AG041689) at the University of Pennsylvania, funded by NIA.

## Alzheimer’s Disease Neuroimaging Initiative (sa000002) data

Data collection and sharing for this project was funded by the Alzheimer’s Disease Neuroimaging Initiative (ADNI) (National Institutes of Health Grant U01 AG024904) and DOD ADNI (Department of Defense award number W81XWH-12-2-0012). ADNI is funded by the National Institute on Aging, the National Institute of Biomedical Imaging and Bioengineering, and through generous contributions from the following: AbbVie, Alzheimer’s Association; Alzheimer’s Drug Discovery Foundation; Araclon Biotech; BioClinica, Inc.; Biogen; Bristol-Myers Squibb Company; CereSpir, Inc.; Cogstate; Eisai Inc.; Elan Pharmaceuticals, Inc.; Eli Lilly and Company; EuroImmun; F. Hoffmann-La Roche Ltd and its affiliated company Genentech, Inc.; Fujirebio; GE Healthcare; IXICO Ltd.; Janssen Alzheimer Immunotherapy Research & Development, LLC.; Johnson & Johnson Pharmaceutical Research & Development LLC.; Lumosity; Lundbeck; Merck & Co., Inc.; Meso Scale Diagnostics, LLC.; NeuroRx Research; Neurotrack Technologies; Novartis Pharmaceuticals Corporation; Pfizer Inc.; Piramal Imaging; Servier; Takeda Pharmaceutical Company; and Transition Therapeutics. The Canadian Institutes of Health Research is providing funds to support ADNI clinical sites in Canada. Private sector contributions are facilitated by the Foundation for the National Institutes of Health (www.fnih.org). The grantee organization is the Northern California Institute for Research and Education, and the study is coordinated by the Alzheimer’s Therapeutic Research Institute at the University of Southern California. ADNI data are disseminated by the Laboratory for Neuro Imaging at the University of Southern California.

Additional information to include in an acknowledgment statement can be found on the LONI site: https://adni.loni.usc.edu/wp-content/uploads/how_to_apply/ADNI_Data_Use_Agreement.pdf.

## Alzheimer’s Disease Genetics Consortium (sa000003) data

Use the following for use of any ADGC generated data:

The Alzheimer’s Disease Genetics Consortium (ADGC) supported sample preparation, sequencing and data processing through NIA grant U01AG032984. Sequencing data generation and harmonization is supported by the Genome Center for Alzheimer’s Disease, U54AG052427, and data sharing is supported by NIAGADS, U24AG041689. Samples from the National Centralized Repository for Alzheimer’s Disease and Related Dementias (NCRAD), which receives government support under a cooperative agreement grant (U24 AG021886) awarded by the National Institute on Aging (NIA), were used in this study. We thank contributors who collected samples used in this study, as well as patients and their families, whose help and participation made this work possible.

For use with the **ADGC-TARCC-WGS** (snd10030) data:

This study was made possible by the Texas Alzheimer’s Research and Care Consortium (TARCC) funded by the state of Texas through the Texas Council on Alzheimer’s Disease and Related Disorders and the Darrell K Royal Texas Alzheimer’s Initiative.

## The Familial Alzheimer Sequencing Project (sa000004) data

This work was supported by grants from the National Institutes of Health (R01AG044546, P01AG003991, RF1AG053303, R01AG058501, U01AG058922, RF1AG058501 and R01AG057777). The recruitment and clinical characterization of research participants at Washington University were supported by NIH P50 AG05681, P01 AG03991, and P01 AG026276. This work was supported by access to equipment made possible by the Hope Center for Neurological Disorders, and the Departments of Neurology and Psychiatry at Washington University School of Medicine.

We thank the contributors who collected samples used in this study, as well as patients and their families, whose help and participation made this work possible. This work was supported by access to equipment made possible by the Hope Center for Neurological Disorders, and the Departments of Neurology and Psychiatry at Washington University School of Medicine

## Charles F. and Joanne Knight Alzheimer’s Disease Research Center (sa000008) data

For use of the ADSP-PHC harmonized phenotypes deposited within dataset, ng00067, use the following statement:

The Memory and Aging Project at the Knight-ADRC (Knight-ADRC), supported by NIH grants R01AG064614, R01AG044546, RF1AG053303, RF1AG058501, U01AG058922 and R01AG064877 to Carlos Cruchaga. The recruitment and clinical characterization of research participants at Washington University was supported by NIH grants P30AG066444, P01AG03991, and P01AG026276. Data collection and sharing for this project was supported by NIH grants RF1AG054080, P30AG066462, R01AG064614 and U01AG052410. This work was supported by access to equipment made possible by the Hope Center for Neurological Disorders, the Neurogenomics and Informatics Center (NGI: https://neurogenomics.wustl.edu/) and the Departments of Neurology and Psychiatry at Washington University School of Medicine.

## Accelerating Medicines Partnership-Alzheimer’s Disease (AMP-AD) (sa000011) data

### Mayo RNAseq Study

- Study data were provided by the following sources: The Mayo Clinic Alzheimer’s Disease Genetic Studies, led by Dr. Nilufer Ertekin-Taner and Dr. Steven G. Younkin, Mayo Clinic, Jacksonville, FL using samples from the Mayo Clinic Study of Aging, the Mayo Clinic Alzheimer’s Disease Research Center, and the Mayo Clinic Brain Bank. Data collection was supported through funding by NIA grants P50 AG016574, R01 AG032990, U01 AG046139, R01 AG018023, U01 AG006576, U01 AG006786, R01 AG025711, R01 AG017216, R01 AG003949, NINDS grant R01 NS080820, CurePSP Foundation, and support from Mayo Foundation. Study data includes samples collected through the Sun Health Research Institute Brain and Body Donation Program of Sun City, Arizona. The Brain and Body Donation Program is supported by the National Institute of Neurological Disorders and Stroke (U24 NS072026 National Brain and Tissue Resource for Parkinson’s Disease and Related Disorders), the National Institute on Aging (P30 AG19610 Arizona Alzheimer’s Disease Core Center), the Arizona Department of Health Services (contract 211002, Arizona Alzheimer’s Research Center), the Arizona Biomedical Research Commission (contracts 4001, 0011, 05-901 and 1001 to the Arizona Parkinson’s Disease Consortium) and the Michael J. Fox Foundation for Parkinson’s Research

### ROSMAP

- We are grateful to the participants in the Religious Order Study, the Memory and Aging Project. This work is supported by the US National Institutes of Health [U01 AG046152, R01 AG043617, R01 AG042210, R01 AG036042, R01 AG036836, R01 AG032990, R01 AG18023, RC2 AG036547, P50 AG016574, U01 ES017155, KL2 RR024151, K25 AG041906-01, R01 AG30146, P30 AG10161, R01 AG17917, R01 AG15819, K08 AG034290, P30 AG10161 and R01 AG11101.

### Mount Sinai Brain Bank (MSBB)

- This work was supported by the grants R01AG046170, RF1AG054014, RF1AG057440 and R01AG057907 from the NIH/National Institute on Aging (NIA). R01AG046170 is a component of the AMP-AD Target Discovery and Preclinical Validation Project. Brain tissue collection and characterization was supported by NIH HHSN271201300031C.

## University of Pittsburg-Kamboh (sa000012) data

This study was supported by the National Institute on Aging (NIA) grants AG030653, AG041718, AG064877 and P30-AG066468.

## NACC Genentech Study (sa000013) data

We would like to thank study participants, their families, and the sample collectors for their invaluable contributions. This research was supported in part by the National Institute on Aging grant U01AG049508 (PI Alison M. Goate). This research was supported in part by Genentech, Inc. (PI Alison M. Goate, Robert R. Graham).

The NACC database is funded by NIA/NIH Grant U01 AG016976. NACC data are contributed by these NIA-funded ADCs: P30 AG013846 (PI Neil Kowall, MD), P50 AG008702 (PI Scott Small, MD), P50 AG025688 (PI Allan Levey, MD, PhD), P30 AG010133 (PI Andrew Saykin, PsyD), P50 AG005146 (PI Marilyn Albert, PhD), P50 AG005134 (PI Bradley Hyman, MD, PhD), P50 AG016574 (PI Ronald Petersen, MD, PhD), P30 AG013854 (PI M. Marsel Mesulam, MD), P30 AG008017 (PI Jeffrey Kaye, MD), P30 AG010161 (PI David Bennett, MD), P30 AG010129 (PI Charles DeCarli, MD), P50 AG016573 (PI Frank LaFerla, PhD), P50 AG005131 (PI Douglas Galasko, MD), P30 AG028383 (PI Linda Van Eldik, PhD), P30 AG010124 (PI John Trojanowski, MD, PhD), P50 AG005142 (PI Helena Chui, MD), P30 AG012300 (PI Roger Rosenberg, MD), P50 AG005136 (PI Thomas Grabowski, MD), P50 AG005681 (PI John Morris, MD), P30 AG028377 (Kathleen Welsh-Bohmer, PhD), and P50 AG008671 (PI Henry Paulson, MD, PhD).

Samples from the National Cell Repository for Alzheimer’s Disease (NCRAD), which receives government support under a cooperative agreement grant (U24 AG21886) awarded by the National Institute on Aging (NIA), were used in this study. We thank contributors who collected samples used in this study, as well as patients and their families, whose help and participation made this work possible.

The Alzheimer’s Disease Genetics Consortium supported the collection of samples used in this study through National Institute on Aging (NIA) grants U01AG032984 and RC2AG036528.

## Cache County Study (sa000014) data

We acknowledge the generous contributions of the Cache County Memory Study participants. Sequencing for this study was funded by RF1AG054052 (PI: John S.K. Kauwe).

## NIH, CurePSP and Tau Consortium PSP WGS (sa000015) data

This project was funded by the NIH grant UG3NS104095 and supported by grants U54NS100693 and U54AG052427. Queen Square Brain Bank is supported by the Reta Lila Weston Institute for Neurological Studies and the Medical Research Council UK. The Mayo Clinic Florida had support from a Morris K. Udall Parkinson’s Disease Research Center of Excellence (NINDS P50 #NS072187), CurePSP and the Tau Consortium. The samples from the University of Pennsylvania are supported by NIA grant P01AG017586.

## CurePSP and Tau Consortium PSP WGS (sa000016) data

This project was funded by the Tau Consortium, Rainwater Charitable Foundation, and CurePSP. It was also supported by NINDS grant U54NS100693 and NIA grants U54NS100693 and U54AG052427. Queen Square Brain Bank is supported by the Reta Lila Weston Institute for Neurological Studies and the Medical Research Council UK. The Mayo Clinic Florida had support from a Morris K. Udall Parkinson’s Disease Research Center of Excellence (NINDS P50 #NS072187), CurePSP and the Tau Consortium. The samples from the University of Pennsylvania are supported by NIA grant P01AG017586. Tissues were received from the Victorian Brain Bank, supported by The Florey Institute of Neuroscience and Mental Health, The Alfred and the Victorian Forensic Institute of Medicine and funded in part by Parkinson’s Victoria and MND Victoria. We are grateful to the Sun Health Research Institute Brain and Body Donation Program of Sun City, Arizona for the provision of human biological materials (or specific description, e.g. brain tissue, cerebrospinal fluid). The Brain and Body Donation Program is supported by the National Institute of Neurological Disorders and Stroke (U24 NS072026 National Brain and Tissue Resource for Parkinson’s Disease and Related Disorders), the National Institute on Aging (P30 AG19610 Arizona Alzheimer’s Disease Core Center), the Arizona Department of Health Services ( contract 211002, Arizona Alzheimer’s Research Center), the Arizona Biomedical Research Commission (contracts 4001, 0011, 05-901 and 1001 to the Arizona Parkinson’s Disease Consortium) and the Michael J. Fox Foundation for Parkinson’s Research. Biomaterial was provided by the Study Group DESCRIBE of theClinical Research of the German Center for Neurodegenerative Diseases (DZNE).

## UCLA Progressive Supranuclear Palsy (sa000017) data

If data are used for a publication, “on behalf of the AL-108-231 investigators” should be included in the authorship list.

## The Diagnostic Assessment of Dementia for the Longitudinal Aging Study of India (LASI-DAD) (sa000019) data

In text: “The Longitudinal Aging Study in India, Diagnostic Assessment of Dementia data is sponsored by the National Institute on Aging (grant numbers R01AG051125 and U01AG065958) and is conducted by the University of Southern California.”

In references: “The Longitudinal Aging Study in India, Diagnostic Assessment of Dementia Study. Produced and distributed by the University of Southern California with funding from the National Institute on Aging (grant numbers R01AG051125 and U01AG065958), Los Angles, CA.”

## Dissecting the Genomic Etiology of non-Mendelian Early-Onset Alzheimer Disease (EOAD) and Related Phenotypes (sa000023) data

This work was supported by the National Institutes of Health (NIH) grant R01AG064614. The ADSP-FUS is supported by U01AG057659.

The National Institutes of Health, National Institute on Aging (NIH-NIA) supported this work through the following grants: ADGC, U01 AG032984, RC2 AG036528; samples from the National Centralized Repository for Alzheimer’s Disease and Related Dementias (NCRAD), which receives government support under a cooperative agreement grant (U24 AG21886) awarded by the National Institute on Aging (NIA), were used in this study. Sequencing data generation and harmonization is supported by the Genome Center for Alzheimer’s Disease, U54AG052427, and data sharing is supported by NIAGADS, U24AG041689. We thank contributors who collected samples used in this study, as well as patients and their families, whose help and participation made this work possible.

NIH grants supported enrollment and data collection for the individual studies including the Alzheimer’s Disease Centers (ADC, P30 AG062429 (PI James Brewer, MD, PhD), P30 AG066468 (PI Oscar Lopez, MD), P30 AG062421 (PI Bradley Hyman, MD, PhD), P30 AG066509 (PI Thomas Grabowski, MD), P30 AG066514 (PI Mary Sano, PhD), P30 AG066530 (PI Helena Chui, MD), P30 AG066507 (PI Marilyn Albert, PhD), P30 AG066444 (PI John Morris, MD), P30 AG066518 (PI Jeffrey Kaye, MD), P30 AG066512 (PI Thomas Wisniewski, MD), P30 AG066462 (PI Scott Small, MD), P30 AG072979 (PI David Wolk, MD), P30 AG072972 (PI Charles DeCarli, MD), P30 AG072976 (PI Andrew Saykin, PsyD), P30 AG072975 (PI David Bennett, MD), P30 AG072978 (PI Neil Kowall, MD), P30 AG072977 (PI Robert Vassar, PhD), P30 AG066519 (PI Frank LaFerla, PhD), P30 AG062677 (PI Ronald Petersen, MD, PhD), P30 AG079280 (PI Eric Reiman, MD), P30 AG062422 (PI Gil Rabinovici, MD), P30 AG066511 (PI Allan Levey, MD, PhD), P30 AG072946 (PI Linda Van Eldik, PhD), P30 AG062715 (PI Sanjay Asthana, MD, FRCP), P30 AG072973 (PI Russell Swerdlow, MD), P30 AG066506 (PI Todd Golde, MD, PhD), P30 AG066508 (PI Stephen Strittmatter, MD, PhD), P30 AG066515 (PI Victor Henderson, MD, MS), P30 AG072947 (PI Suzanne Craft, PhD), P30 AG072931 (PI Henry Paulson, MD, PhD), P30 AG066546 (PI Sudha Seshadri, MD), P20 AG068024 (PI Erik Roberson, MD, PhD), P20 AG068053 (PI Justin Miller, PhD), P20 AG068077 (PI Gary Rosenberg, MD), P20 AG068082 (PI Angela Jefferson, PhD), P30 AG072958 (PI Heather Whitson, MD), P30 AG072959 (PI James Leverenz, MD). The Miami ascertainment and research were supported in part through: RF1AG054080, R01AG027944, R01AG019085, R01AG028786-02, RC2AG036528. The Columbia ascertainment and research were supported in part through: R37AG015473 and U24AG056270. The University of Washington ascertainment and research were supported in part through R01AG044546, RF1AG053303, RF1AG058501, U01AG058922 and R01AG064877.

## Code availability

- VCPA code https://bitbucket.org/NIAGADS/vcpa-pipeline/src/master/
- SV related code

`o` https://github.com/DecodeGenetics/svimmer, https://github.com/DecodeGenetics/graphtyper
- QC code https://bitbucket.org/Taha_Iqbal_UPenn/gcad-vcf-qc_public/

## Data availability

- Complete list of files showing the NIAGADS accession number together with a description of the files is available at Supplementary Table S5.
- NIAGADS DSS is accessible at https://dss.niagads.org/
- VariXam is accessible at https://varixam.niagads.org/
- GenomicsDB is accessible at https://www.niagads.org/genomics/app

## Notes

### Competing Interest Statement

Timothy Hohman serves in the Scientific Advisory Board for Vivid Genomics, and Deputy Editor for Alzheimer's & Dementia: TRCI.

### Author Declarations

In this manuscript, all data are from NIAGADS. NIAGADS contains both individual level and aggregated/summary data. All individual level data has been de-identified and NIAGADS users are prohibited from trying to reidentify individuals as part of the code of conduct they sign when applying for data within NIAGADS. Open-access data can be freely downloaded from the NIAGADS website. Restricted-access data are available through the NIAGADS Data Sharing Service (dss.niagads.org) and require an approved data access request to ensure compliance with privacy and ethical guidelines and in accordance with the terms outlined by the submitting Institutional Review Boards (IRBs) and the consent provided by research participants. For detailed instructions on accessing specific datasets, including open and restricted data, please visit the NIAGADS website at www.niagads.org

