## Supplementary Figures and Methods for "Alzheimer’s Disease Sequencing Project Release 4 Whole Genome Sequencing Dataset"

**Supplementary Figure 1 – ADSP Data flow: from receipt to distribution.**

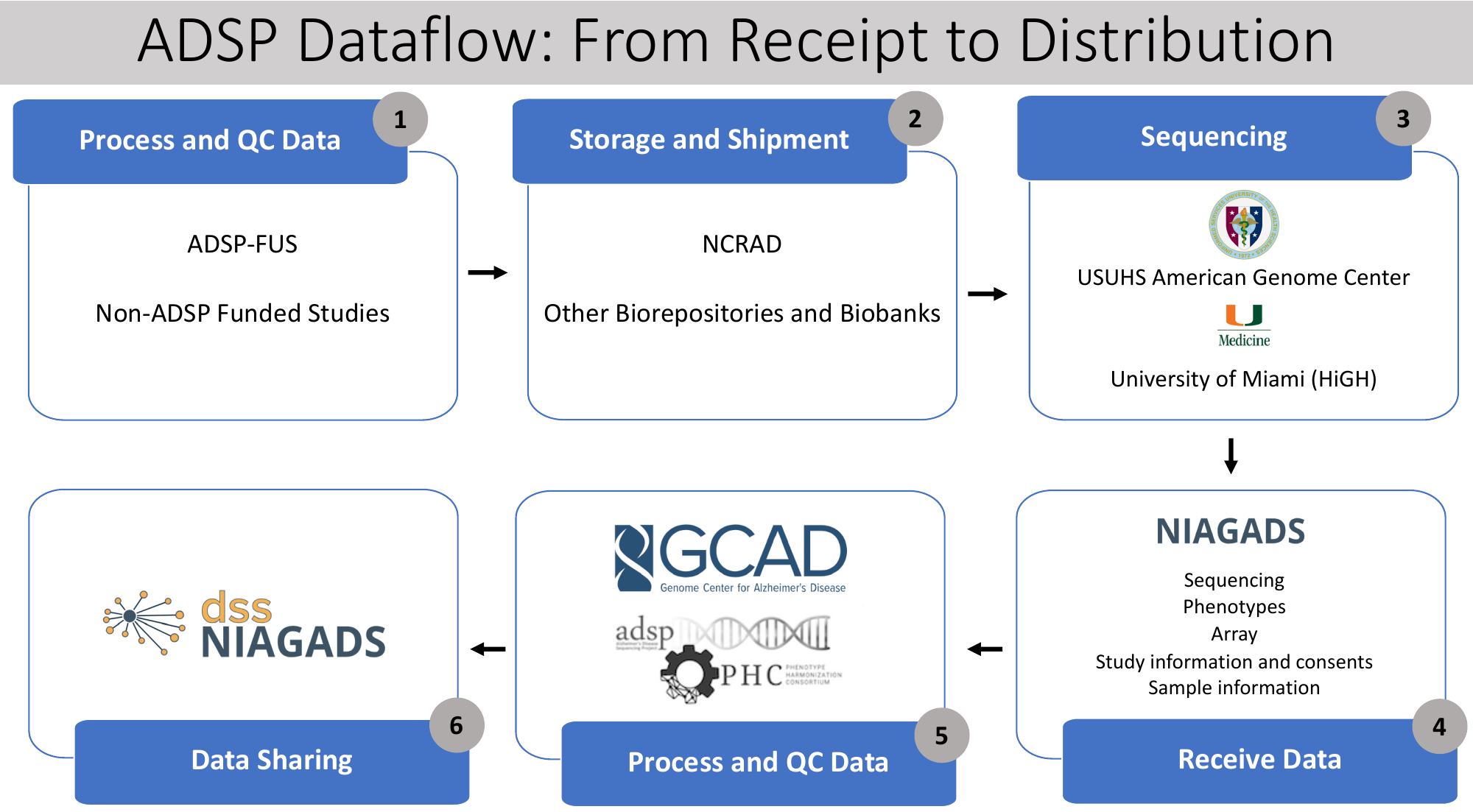

**Supplementary Figure 2a –** Number of indels called per sample in each reported ethnic group. Line in each displayed boxplot denotes the mean value where each dot is a sample. **Supplementary Figure 2b –** Number of SNVs across reported ethnic groups, broken down by sequencing platform (columns) and PCR protocol (rows).

**(a)**

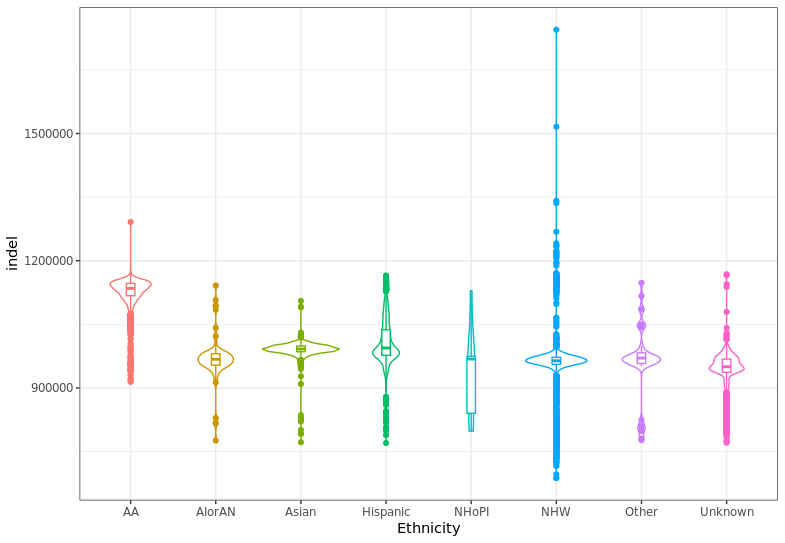

**(b)**

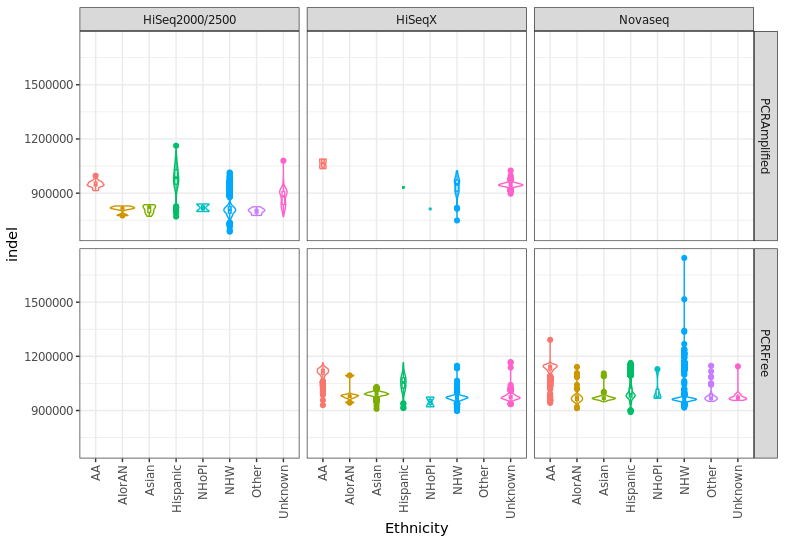

**Supplementary Figure 3 – Breakdown of ethnicity across ADSP R3 and R4 data.**

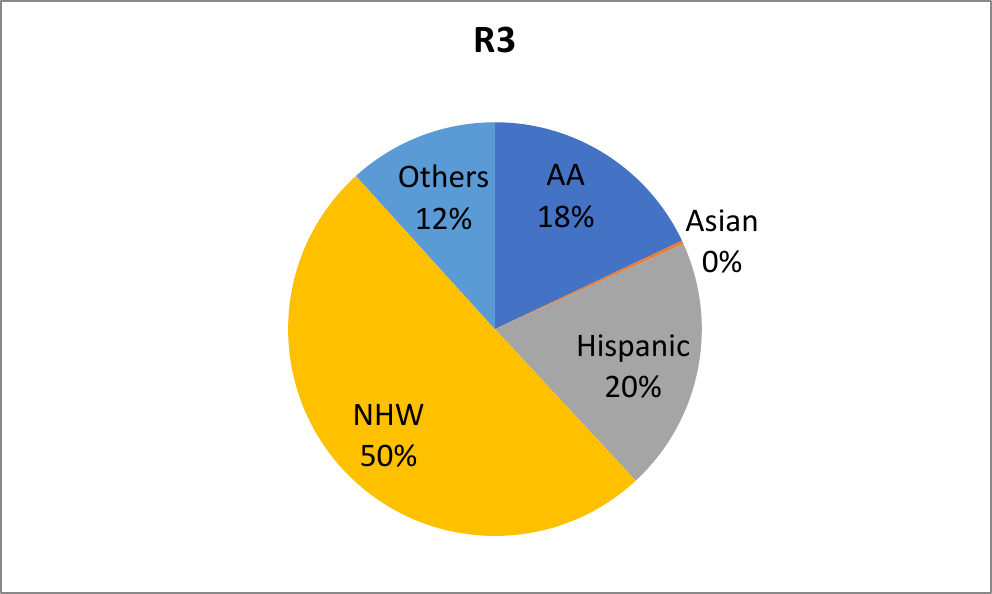

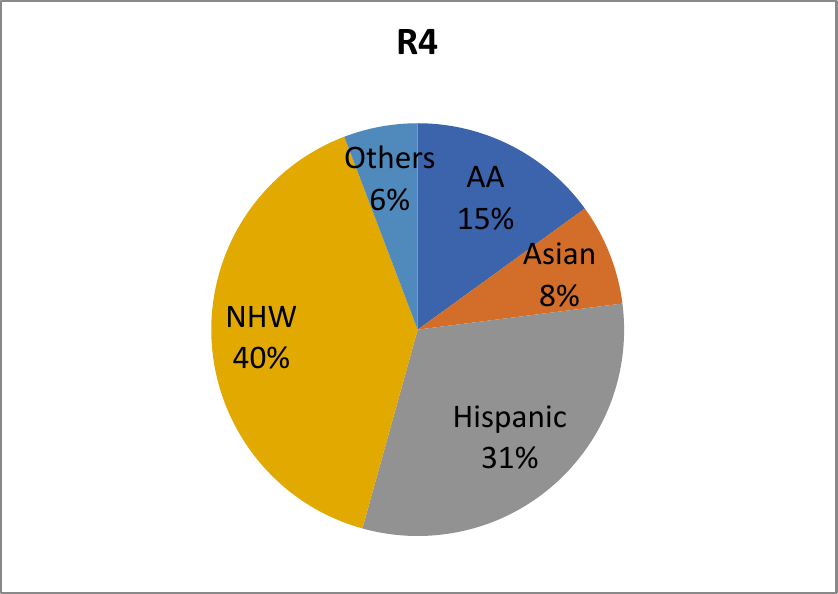

**Supplementary Figure 4 – Dataflow and pipelines for ADSP R4 data.** The official ADSP pipeline, VCPA, accepts genomes in FASTQs/BAMs/CRAMs format (grey boxes). CRAMs and gVCFs are the outputs per individual genome (blue boxes). gVCFs across samples are inputs for joint genotyping, resulting in joint called pVCFs and other VCFs with QCed info or smaller file sizes (green boxes). Individual CRAMs are used for SV calls and subsequently get joint genotyped into a pVCF (yellow boxes). Boxes marked with “star” are available in NIAGADS DSS.

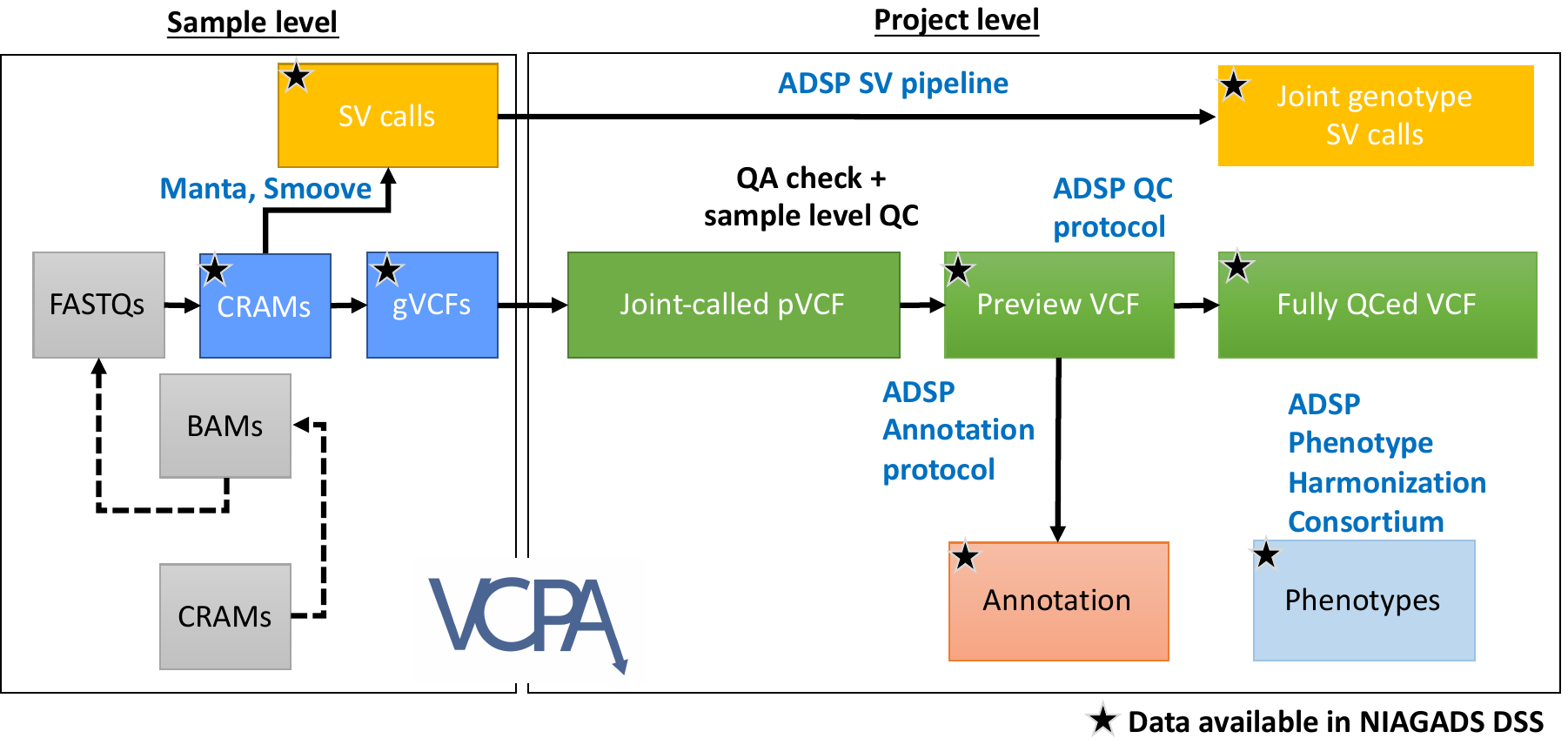

**Supplementary Figure 5 – Sample level QC process.**

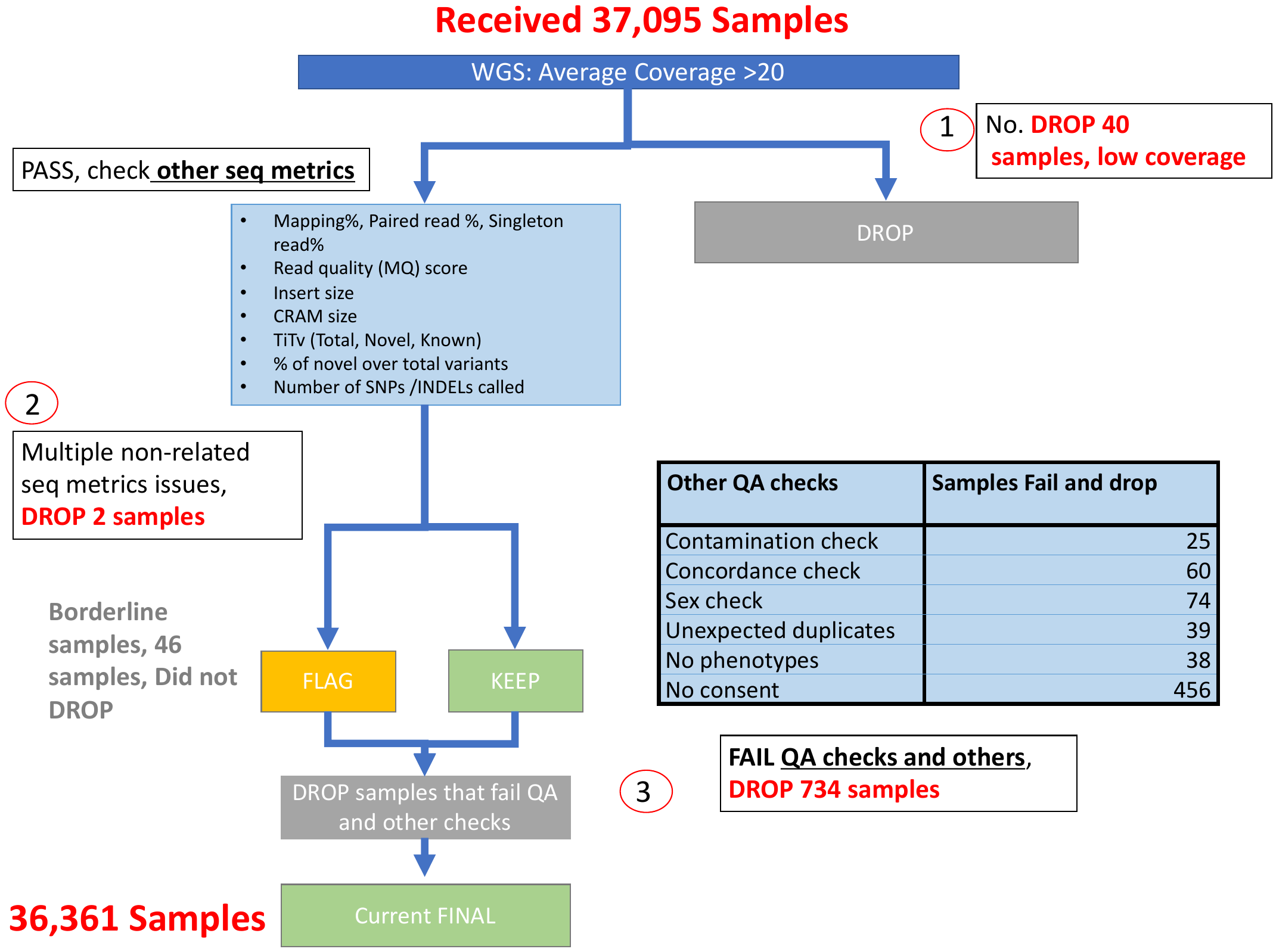

**Supplementary Methods**

**Cohorts description**

### Adult Changes in Thought (ACT)

The Adult Changes in Thought (ACT) study is a longitudinal prospective cohort study that began in 1994.  Participants are randomly selected from Seattle area members of Group Health aged 65 years or older.  Participants are cognitively intact at study enrollment, defined operationally as a CASI (Cognitive Abilities Screening Instrument) score of >85 or consensus diagnosis of “not demented” following comprehensive neurological and neuropsychological assessment.  Incident cases of dementia are identified with the same 2-stage sampling scheme, where all participants with CASI scores <86 are evaluated with a comprehensive neurological and neuropsychological assessment, and all results are considered at a consensus conference.  Autopsy is also available for this collection; autopsy consent rates are about 25% of the cohort.  Genomic DNA from blood and/or brain tissue is available from this collection.  This study includes 4690 subjects across three sub-cohorts.

**Website:**

<https://www.maelstrom-research.org/mica/individual-study/act>

### Alzheimer’s Disease Neuroimaging Initiative (ADNI)

ADNI is a global research study that actively supports the investigation and development of treatments that slow or stop the progression of AD. In this multisite longitudinal study, researchers at 63 sites in the US and Canada track the progression of AD in the human brain with clinical, imaging, genetic and biospecimen biomarkers through the process of normal aging, early mild cognitive impairment (EMCI), and late mild cognitive impairment (LMCI) to dementia or AD. Participants undergo a series of initial tests that are repeated at intervals over subsequent years, including a clinical evaluation, neuropsychological tests, genetic testing, lumbar puncture, and MRI and PET scans. The overall goal of ADNI is to validate biomarkers for use in Alzheimer’ s disease clinical treatment trials. 1338 cases and 483 controls are included in this study across 4 phases.

### Amish Protective Variant Study

The Amish Protective Variant Study, based at Case Western Reserve University and the University of Miami (Cummings et al. 2012; D’Aoust et al. 2015), is being conducted in the Indiana and Ohio Old Order Amish (OOA) population. The OOA, an isolated founder population originating from emigration of German and Swiss Anabaptists to the U.S. in the 1700’s and 1800’s has been examined in multiple genetic studies of complex diseases including AD, age-related macular degeneration, and successful aging. With fewer than 1,000 founders and self-imposed cultural and religious isolation, the introduction of genetic variation among the OOA has been significantly restricted. Anecdotally, their agrarian lifestyle and firm behavioral norms likely have reduced variation in environmental exposures. Further, the OOA are an excellent population for genetic studies in that they have large and stable pedigrees and low variability in lifestyle factors.

Individuals included in this study have been recruited over the past 20 years for multiple studies of AD or dementia, age-related macular degeneration, and successful aging. For all these studies, the primary criteria for enrollment included being age 50 or older (AMD), age 60 or older (AD), or age 80 or older (successful aging), being part of the Amish community, and being of Amish descent. Recruitment primarily included community-based home visits. Participants were recruited from Amish families living in Holmes County, Ohio and Elkhart, LaGrange, and Adams Counties, Indiana.

Nuclear family pedigrees are provided for convenience. However, as a founder population, these Amish individuals have, on average, a genetic relationship between the equivalent of second and third cousins. Thus additional, more complex relationships exist across the nuclear pedigrees.

### Cache County Study (CCS)

The Cache County Study on Memory Health and Aging (CCS) was initiated in 1994 to investigate the association of APOE genotype and environmental exposures on cognitive function and dementia. This cohort of 5092 Cache County, Utah, residents (90% of those aged 65 years or older in 1994), has been followed continuously for over 15 years, with four triennial waves of data collection and additional clinical assessments for those at high-risk for dementia. DNA samples were obtained from 97.6% of participants. The Cache County population is exceptionally long-lived and ranked number one in life expectancy among all counties in the 1990 US Census. All but one of the members of the CCS have been linked to the UPDB and their extended genealogies are known. This population was the source of most of the Centre d’Etude du Polymorphisme Humain (CEPH) families that have been used to represent Caucasians in many genetic studies worldwide, including the HapMap project. Recent analyses confirm that these data are representative of the general European-American population. For this study, we needed both AD cases and resilient individuals identified in the same pedigrees.” (Ridge, P.G., Karch, C.M., Hsu, S. et al. Linkage, whole genome sequence, and biological data implicate variants in RAB10 in Alzheimer’s disease resilience. Genome Med 9, 100 (2017). <https://doi.org/10.1186/s13073-017-0486-1>)

### Case Western Reserve University (CWRU) Autopsy

The Case Western Reserve University (CWRU) Autopsy Cohort was an Alzheimer’s Disease Research Center (ADRC) clinic-based sample. Subjects included in the Case Western Reserve Autopsy cohort include individuals who were participants in the Brain Health and Memory Center Research Brain Donation Protocol at University Hospitals Cleveland Medical Center in Cleveland, OH. The target population is patients who have degenerative disorders of the central nervous system. Recruitment of potential subjects is primarily based on referrals from health care providers. In addition, individuals who learn of the program through other means may participate. This is an autopsy-based study designed to analyze post-mortem brain tissue. Medical records are also obtained as available to help correlate brain behavior relationships.

Participants were classified into clinical categories (Alzheimer’s disease, Controls, and other dementia (ADRD) based on description of the brain gross examination and neuropath microscopic description. Cases consisted of individuals who were diagnosed as AD. Controls consisted of individuals who had insufficient findings of neurofibrillary tangles or BRAAK stage I/II or essentially normal brain for age. Other (ADRD) consisted of individuals with diagnosis not being AD or Control.

### Case Western Reserve University (CWRU) Rapid Decline

The Case Western Reserve University (CWRU) Rapid Decline Cohort (Cohen et al. 2015) represents patients with rapid decline. Subjects included in the Case Western Reserve Rapid Decline dataset include individuals who were initially suspected of having Creutzfelt-Jakob disease (CJD). The Brains of these individuals were obtained by the National Prion Disease Pathology Surveillance Center (NPDPSC) for testing and confirmation of CJD. Samples are received from across the United States. After testing, these samples were determined not to have CJD, but to have pathology consistent with Alzheimer disease. The usual progression of CJD is quite rapid, so that the vast majority of these individuals progressed from onset to death in less than 3 years. This is an autopsy-based study designed to analyze post-mortem brain tissue. Medical records are obtained when available to help correlate brain behavior relationships.

Participants were classified as either Alzheimer’s disease or another dementia (ADRD; Lewy body Dementia, Frontotemporal dementia) based on description of the brain gross examination and neuropath microscopic description.

### Cuban American Alzheimer’s Disease Initiative (CuAADI)

The Cuban American Alzheimer’s Disease Initiative (CuAADI) is a convenience sample ascertained through community outreach to Alzheimer’s and adult day care centers in Southern Florida, lay conferences, and Neurology and Memory Disorders clinics. Eligibility was based on self-reported Cuban heritage. The participants were recruited in South Florida which is home to the largest number of Cubans in the US. Most of our participants have been living in South Florida since they moved from Cuba in the last 20 to 45 years.

Participants were ascertained and evaluated through community centers, referrals from University of Miami memory clinics and adult day care centers. Some participants were evaluated in their homes. All participants greater than 60 years of age underwent a standard clinical evaluation consisting of a medical and family history interview, neuropsychological testing, behavioral and emotional assessments, and functional measures. Venous blood samples (or saliva samples when needed) were collected on all participants. All assessments were conducted in the preferred language of the participant or knowledgeable informant.

### Erasmus Rucphen Family (ERF)

A family-based cohort study that is embedded in the Genetic Research in Isolated Populations (GRIP) program in the South West of the Netherlands. The aim of this program was to identify genetic risk factors in the development of complex disorders. For the Erasmus Rucphen Family Study (ERF), 22 families that had at least five children baptized in the community church between 1850-1900 were identified with the help of genealogical records. All living descendants of these couples and their spouses were invited to take part in the study. Data collection started in June 2002 and was finished in February 2005 (n=2065).

**Website:**

<https://www.neurodegenerationresearch.eu/cohort/erasmus-rucphen-family-study/>

### Estudio Familiar de Influencia Genetica en Alzheimer (EFIGA)

Estudio Familiar de Influencia Genetica en Alzheimer (EFIGA) included 683 at-risk family members from 242 AD-affected families of Caribbean Hispanic descent. These families have 2 or more individuals affected with Alzheimer’s disease. A system of recruitment was also set up in the Dominican Republic with the help of several local physicians, including the president of the Dominican Society of Geriatrics and Gerontology. All affected and unaffected family members are evaluated in person both in the Dominican Republic and New York. A case was defined as any individual meeting NINCDS-ADRDA criteria for probable or possible LOAD. The Clinical Dementia Rating was used to rate the severity of dementia. Brain imaging and other laboratory study results were reviewed, when available, to ensure full implementation of the NINCDS-ADRDA criteria.

Once patients with LOAD were identified, their illnesses were documented with standardized neurological and neuropsychological evaluations. Structured family history interviews were then conducted with available family members to determine whether patients had living siblings or relatives with the disease. Medical and neurological examinations were completed for all family members. Brains of participants with dementia and history of stroke were administered magnetic resonance imaging scans to exclude patients with comorbid cerebrovascular disease. DNA samples and cell lines are stored for all participating individuals.

The goal of this study is to root out genetic variants that increase late onset Alzheimer disease risk in this ethnic group. This study was initiated in 1998 and recruited subjects from the Taub Institute for Research on Alzheimer’s Disease and the Aging Brain in New York as well as from clinics in the Dominican Republic.

**Website:**

<http://www.cumc.columbia.edu/adrc/investigators>

### Harmonized Diagnostic Assessment of Dementia for the Longitudinal Aging Study of India (LASI-DAD)

LASI-DAD is an in-depth study of late-life cognition and dementia. It draws a sub-sample of 4,000+ LASI respondents aged 60 and older and administers in-depth cognitive tests and informant interviews. LASI is an ongoing cohort study of 72,000 community-residing older adults aged 45 and older and their spouses regardless of age, with an oversample of persons aged 65 and above.The study design involves following all individuals who move into institutions, although that number is expected to be very small in India. Using the Census as the sampling frame, LASI is representative of both the nation as a whole and each state and union territory. Due to administrative and linguistic considerations, the fieldwork for the main LASI unfolded in three phases, and therefore the LASI-DAD fieldwork was also carried out in phases, recruiting most respondents about 6 – 7 months after the core LASI interview.

**Website:**

<https://lasi-dad.org/>

### Hillblom Aging Network (HAN)

Participants were enrolled in the Hillblom Aging Network at the University of California, San Francisco (UCSF) Memory and Aging Center. All participants underwent comprehensive neurobehavioral evaluations and met the following inclusionary criteria at baseline: 1) clinically normal based on consensus conference with a neurologist and board-certified neuropsychologist; 2) no history of neurological disorder known to impact cognition (e.g., epilepsy, stroke); and 3) functionally intact as defined by an informant-obtained CDR global score of 0 ([Morris, 1993](https://www.ncbi.nlm.nih.gov/pmc/articles/PMC7839841/#R49)). More specifically, the determination of clinically normal by consensus conference involved ruling out the presence of mild cognitive impairment, dementia, or any other neurological condition resulting in cognitive, behavioral, motor, or functional decline (e.g., Parkinson’s disease), according to widely used diagnostic criteria (e.g., [Albert et al., 2011](https://www.ncbi.nlm.nih.gov/pmc/articles/PMC7839841/#R1); [Armstrong et al., 2013](https://www.ncbi.nlm.nih.gov/pmc/articles/PMC7839841/#R4); [Gorno-Tempini et al., 2011](https://www.ncbi.nlm.nih.gov/pmc/articles/PMC7839841/" \l "R24" \t "_blank); [Höglinger et al., 2017](https://www.ncbi.nlm.nih.gov/pmc/articles/PMC7839841/" \l "R30" \t "_blank); [McKeith et al., 2017](https://www.ncbi.nlm.nih.gov/pmc/articles/PMC7839841/" \l "R45" \t "_blank); [McKhann et al., 2011](https://www.ncbi.nlm.nih.gov/pmc/articles/PMC7839841/" \l "R46" \t "_blank); [Postuma et al., 2015](https://www.ncbi.nlm.nih.gov/pmc/articles/PMC7839841/" \l "R55" \t "_blank); [Rascovsky et al., 2011](https://www.ncbi.nlm.nih.gov/pmc/articles/PMC7839841/" \l "R56" \t "_blank)). Three main sources of information were considered by the neurologist and neuropsychologist during the diagnostic conference. First, participants underwent a thorough evaluation with the neurologist that involved a comprehensive neurological examination, clinical interview, and review of systems. Second, neuroimaging (structural MRI) was reviewed to screen out gross brain pathology with potential to negatively impact cognition (e.g., tumor). Third, participants completed a battery of neuropsychological tests to objectively assess major domains of cognitive function, including attention, executive functioning, memory, language, and visuospatial skills. Cognitive impairment was defined by the presence of subjective cognitive decline, as reported by the participant or informant, together with objective performance on neuropsychological testing that was below expectation given the participant’s age and level of premorbid functioning ([Albert et al., 2011](https://www.ncbi.nlm.nih.gov/pmc/articles/PMC7839841/#R1)). In making the determination of clinically normal, emphasis was placed on ruling out any declines in the participant’s ability to perform everyday tasks due to cognitive changes.

University of California San Francisco Alzheimer’s Disease Research Center under grant P30AG062422;

Larry L. Hillblom Network under Grant 2014-A-004-NET;

R01AG032289 (PI: JK);

R01AG048234 (PI: JK)

Espeland MA, Yassine H, Hayden KD, Hugenschmidt C, Bennett WL, Chao A, Neiberg R, Kahn SE, Luchsinger JA; Action for Health in Diabetes (Look AHEAD) Research Group. Sex-related differences in cognitive trajectories in older individuals with type 2 diabetes and overweight or obesity. Alzheimers Dement (N Y). 2021 Apr 9;7(1):e12160. doi: 10.1002/trc2.12160. PMID: [33860069](https://pubmed.ncbi.nlm.nih.gov/33860069/); PMCID: PMC8033410.

Casaletto KB, Elahi FM, Staffaroni AM, Walters S, Contreras WR, Wolf A, Dubal D, Miller B, Yaffe K, Kramer JH. Cognitive aging is not created equally: differentiating unique cognitive phenotypes in “normal” adults. Neurobiol Aging. 2019 May;77:13-19. doi: 10.1016/j.neurobiolaging.2019.01.007. Epub 2019 Jan 24. PMID: 30772736; PMCID: PMC6486874.

Staffaroni AM, Brown JA, Casaletto KB, Elahi FM, Deng J, Neuhaus J, Cobigo Y, Mumford PS, Walters S, Saloner R, Karydas A, Coppola G, Rosen HJ, Miller BL, Seeley WW, Kramer JH. The Longitudinal Trajectory of Default Mode Network Connectivity in Healthy Older Adults Varies As a Function of Age and Is Associated with Changes in Episodic Memory and Processing Speed. J Neurosci. 2018 Mar 14;38(11):2809-2817. doi: 10.1523/JNEUROSCI.3067-17.2018. Epub 2018 Feb 13. PMID: 29440553; PMCID: PMC5852659.

Yokoyama JS, Sturm VE, Bonham LW, Klein E, Arfanakis K, Yu L, Coppola G, Kramer JH, Bennett DA, Miller BL, Dubal DB. Variation in longevity gene KLOTHO is associated with greater cortical volumes. Ann Clin Transl Neurol. 2015 Mar;2(3):215-30. doi: 10.1002/acn3.161. Epub 2015 Jan 26. PMID: 25815349; PMCID: PMC4369272.

### Indianapolis-Ibadan (IIAA/IIBD)

The Indianapolis-Ibadan Dementia Project, established in 1991, is a longitudinal prospective population-based comparative epidemiological study of the prevalence and incidence rates and risk factors for Alzheimer’s disease and other age associated dementias. Enrollment of community-dwelling elderly (age>65 years) African Americans living in Indianapolis and Yoruba living in Ibadan, Nigeria employed the same research design, methods, and investigators (see study description at <https://iidpportal.medicine.iu.edu/>). The first enrollment wave began in 1992 and participants were followed every 2 to 3 years.

Participants were ascertained and evaluated through community centers, clinical and hospital settings as well in community centers and at home. All participants greater than 65 years of age who agreed to participate were screened using measures. Those who failed the screen underwent a more comprehensive clinical evaluation. Medical and family history interview, neuropsychological testing, behavioral and emotional assessments, and functional measures, including collateral informant report, were available for all most participants. Venous blood samples (were collected on all participants. All assessments were conducted in the preferred language of the participant or knowledgeable informant. Finally, all participants were adjudicated by a clinical consensus panel and were classified according to various criteria in place at the time of the clinical data collection Details on diagnosis criteria and process were described in Hendrie et al JAMA 2001.

**Website:**

<https://iidpportal.medicine.iu.edu/>

### Knight Alzheimer’s Disease Research Center (KGAD)

The search for novel risk factors for Alzheimer disease relies on access to accurate and deeply phenotyped datasets. The Memory and Aging Project at the Knight-ADRC (Knight ADRC-MAP) collects plasma, CSF, fibroblast, neuroimaging, clinical and cognition data longitudinally and autopsied brain samples. We are using multi-tissue (brain, CSF and plasma) multi-omic data (genetics, epigenomics, transcriptomics, proteomics and metabolomics) to identify novel risk and protective variants, create new prediction models and identify drug targets. The study cohort includes MAP participants from the Knight-ADRC at Washington University in St. Louis (MO). MAP participants have to be at least 65 years old and have no memory problems or mild dementia at the time of enrollment.  There is no age at onset criteria for this cohort. Cases had to have a CDR >=0.5 whereas controls had to have a CDR=0 at last assessment.  AD definition is based on a combination of both clinical and pathological information if available. Pathologic diagnosis will overrule clinical diagnosis.  Participants are Non-Hispanic white from North America (95%) and African American (5%). Autopsy information was provided if available, but it is not a requirement for enrollment.

**Website:**

<https://knightadrc.wustl.edu/>

### LonGenity

The LonGenity study at Albert Einstein College of Medicine has been recruiting community dwelling Ashkenazi Jewish seniors aged 65 or older in the United States since 2008. Offspring of Parents with Exceptional Longevity (OPEL), defined by having at least one parent who lived to age 95 or older and Offspring of Parents with Usual Survival (OPUS), defined by having neither parent survived to age 95 are being followed annually in this longitudinal study. The goal of this study is to search for longevity genes that may act to slow the aging process and/or protect from age-related diseases. Participants undergo comprehensive cognitive testing, physical performance assessments, and complete medical and family history questionnaires at annual visits. Blood samples are collected biennially and are used for DNA analysis. Participants selected for this sub-study were either (1) age ³70, carriers of APOe4/e4 genotype, and exhibited normal cognitive function or (2) were age ³80, carriers of APOe3/e4 genotype, and exhibited normal cognitive function. Cognitive function was evaluated annually with comprehensive neurocognitive test battery.

Funding:

Grants from the National Institutes of Health R01AG042188, R01AG044829, R01AG046949, R01AG057909, R01AG061155, P30AG038072, the Einstein-Paul Glenn Foundation for Medical Research Center for the Biology of Human Aging.

### Longevity Genes Project (LGP)

The Longevity Genes Project (LGP), established in 1998 at Albert Einstein College of Medicine, recruits Ashkenazi Jewish centenarians (age 95 and older) who are in general good health at age 95, offspring of centenarians, and spouses of offspring in the Eastern United States. The goal of this cross-sectional study is to identify longevity genes that help to slow the aging process and/or protect from age-related diseases. Participants undergo a physical examination (including physical measurements and mini mental state examination (MMSE)), complete a series of questionnaires (including medical and family history, physical activity, etc.), as well as a blood draw or cheek swab collection for DNA analysis. Participants selected for this sub-study were living in the community and were either (1) age ³70, carriers of APOe4/e4 genotype, and exhibited normal cognitive function or (2) were age ³80, carriers of APOe3/e4 genotype, and exhibited normal cognitive function. For individuals age 95 and older, normal cognitive function was defined as full MMSE score >22 or blind MMSE score ³16. Fo individuals age <95, normal cognition was defined as MMSE >25.

Funding:

Grants from the National Institutes of Health R01AG042188, R01AG044829, R01AG046949, R01AG057909, R01AG061155, P30AG038072, the Einstein-Paul Glenn Foundation for Medical Research Center for the Biology of Human Aging.

### Mayo Clinic (MAYO)

All 248 cases and 98 controls consisted of Caucasian subjects from the United States ascertained at the Mayo Clinic. All subjects were diagnosed by a neurologist at the Mayo Clinic in Jacksonville, Florida or Rochester, Minnesota. The neurologist confirmed a Clinical Dementia Rating score of 0 for all controls; cases had diagnoses of possible or probable AD made according to NINCDS-ADRDA criteria. Autopsy-confirmed samples came from the brain bank at the Mayo Clinic in Jacksonville, FL and were evaluated by a single neuropathologist. In clinically-identified cases, the diagnosis of definite AD was made according to NINCDS-ADRDA criteria.

### Mexican Health and Aging Study (MHAS)

This is a national longitudinal study of adults 50 years and older in Mexico.The baseline survey, with national and urban/rural representation of adults born in 1951 or earlier, was conducted in 2001 with follow-up interviews in 2003, 2012, 2015, and 2018. A new sample of adults born between 1952-1962 was added in 2012. Similarly, in 2018 a new cohort of adults born between 1963 and 1968 was added to refresh the sample.

The study is a collaborative effort among researchers from the University of Texas Medical Branch (UTMB), the Instituto Nacional de Estadística y Geografía (INEGI, Mexico), the University of Wisconsin, the Instituto Nacional de Geriatría (INGER, Mexico), the Instituto Nacional de Salud Pública (INSP, Mexico), and University of California Los Angeles (UCLA). The MHAS is partly supported by the National Institutes of Health/National Institute on Aging (R01AG018016, R Wong, PI) in the United States and the Instituto Nacional de Estadística y Geografía (INEGI) in Mexico.

Cohort description taken from the [MHAS website](http://www.mhasweb.org/index.aspx), July 29, 2022.

**Website:**

<http://www.mhasweb.org/index.aspx>

### Mount Sinai Brain Bank (MSBB)

Human brains were accessed from the Mount Sinai/JJ Peters VA Medical Center Brain Bank (MSBB–Mount Sinai NIH Neurobiobank) cohort, which holds over 2,040 well-characterized brains. This cohort was assembled after applying stringent inclusion/exclusion criteria and represents the full spectrum of cognitive and neuropathological disease severity in the absence of discernable non-AD neuropathology. For each sample, neuropathological assessment was performed according to the Consortium to Establish a Registry for Alzheimer’s Disease (CERAD) protocol and included assessment by hematoxylin and eosin, modified Bielschowski, modified thioflavin S, and anti-β amyloid (4G8), anti-tau (AD2) and anti-ubiquitin. A Braak AD-staging score for progression of neurofibrillary neuropathology was assigned to each case. Quantitative data regarding the mean of the density of neuritic plaques in the middle frontal gyrus, orbital frontal cortex, superior temporal gyrus, inferior parietal cortex and calcarine cortex were also collected. Clinical dementia rating scale (CDR) was conducted for assessment of dementia and cognitive status. (Wang, M., Beckmann, N., Roussos, P. et al. The Mount Sinai cohort of large-scale genomic, transcriptomic and proteomic data in Alzheimer’s disease. Sci Data 5, 180185 (2018). <https://doi.org/10.1038/sdata.2018.185>)

### National Cell Repository for Alzheimer’s Disease Family (NCRAD Family)

NCRAD family cohort was started in 1990 and consists of families with two or more members with early or late onset AD and related dementias.  This collection maintains DNA and cell lines on affected family members and unaffected relatives (typically over age 60).  These families are not evaluated in person and all clinical information is obtained through medical record review. Therefore, data is limited to the following: family history; demographic data; medical records on the evaluation; diagnosis and treatment of symptomatic subjects; telephone cognitive battery; neuropathological findings when available. This is a longitudinal study with autopsy available to all participants.  Genomic DNA, Cell Line DNA, Lymphoblastoid Cell Lines (LCLs), and PBMCs are available.

**Website:**

<https://ncrad.iu.edu/accessing_data.html>

### National Institute on Aging Late Onset of Alzheimer’s Disease Family (NIA-LOAD)

The LOAD collection is a longitudinal, multi-center late onset AD sibling genetics initiative.  This NIA-funded study began in 2002 and maintains DNA and cell lines on families with 2 or more siblings with AD (at least one probable or confirmed AD) (n=5291).  A third family member who is either affected or unaffected is also required.  These individuals are evaluated in person and/or over the phone. A minimum dataset is collected for each person in the family. Definite AD is defined by established neuropathological criteria (and confirmed by autopsy). Probable or possible AD is defined according to NINCDS-ADRDA criteria.  Autopsy is offered to all subjects.

**Website:**

<https://www.alz.washington.edu/WEB/researcher_home.html>

### NIA Alzheimer Disease Research Centers (ADRC)

The NIA ADRC cohort included subjects ascertained and evaluated by the clinical and neuropathology cores of the 32 NIA-funded ADRCs. Data collection is coordinated by the National Alzheimer’s Coordinating Center (NACC). NACC coordinates collection of phenotype data from the 32 ADRCs, cleans all data, coordinates implementation of definitions of AD cases and controls, and coordinates collection of samples. The ADRC cohort consists of autopsy-confirmed and clinically-confirmed AD cases, and cognitively normal elders (CNEs) with complete neuropathology data who were older than 60 years at age of death, and living CNEs evaluated using the Uniform dataset (UDS) protocol who were documented to not have mild cognitive impairment (MCI) and were between 60 and 100 years of age at assessment.

Based on the data collected by NACC, the ADGC Neuropathology Core Leaders Subcommittee derived inclusion and exclusion criteria for AD and control samples. All autopsied subjects were age ≥ 60 years at death. AD cases were demented according to DSM-IV criteria or Clinical Dementia Rating (CDR) ≥1. Neuropathologic stratification of cases followed NIA/Reagan criteria explicitly, or used a similar approach when NIA/Reagan criteria were coded as not done, missing, or unknown. Cases were intermediate or high likelihood by NIA/Reagan criteria with moderate to frequent amyloid plaques and neurofibrillary tangle (NFT) Braak stage of III-VI. Persons with Down syndrome, non-AD tauopathies and synucleinopathies were excluded. All autopsied controls had a clinical evaluation within two years of death.

Controls did not meet DSM-IV criteria for dementia, did not have a diagnosis of mild cognitive impairment (MCI), and had a CDR of 0, if performed. Controls did not meet or were low-likelihood AD by NIA/Reagan criteria, had sparse or no amyloid plaques, and a Braak NFT stage of 0 – II. ADRCs sent frozen tissue from autopsied subjects and DNA samples from some autopsied subjects and from living subjects to the ADRCs to the National Cell Repository for Alzheimer’s Disease (NCRAD). DNA was prepared by NCRAD for genotyping and sent to the genotyping site at Children’s Hospital of Philadelphia. ADRC samples were genotyped and analyzed in separate batches.

### Northern Manhattan Study (NOMAS)

This is a research study of stroke and stroke risk factors among the multi-ethnic community of Northern Manhattan, New York. The study is a collaboration between the Department of Neurology at University of Miami and the Neurological Institute at Columbia University.

NOMAS is a NINDS-funded study of the population of Washington Heights in Northern Manhattan. The ongoing study, which began in 1990, is now a collaboration between the Department of Neurology at University of Miami and the Neurological Institute at Columbia University. The study’s interdisciplinary team of doctors and researchers has enrolled over 4,400 people from the community, some of whom have suffered a stroke or related neurological diseases (Sacco et al. 2004).

**Website:**

<https://northernmanhattanstudy.org/>

### Peru Alzheimer’s Disease Initiative (PeADI)

PeADI is being conducted at several sites in Peru including Lima (Marca-Ysabel et al. 2021). The Mestizo population living in Lima, the capital city, is comprised of ~76% of Amerindian, 11% Southern European, and 3% Northern African ancestries, and a smaller proportion of western central Africa, western central Europe, Finland, northern Siberia and Jewish diaspora. Cases will be clinic based and controls will be clinic and community based.

Participants were ascertained for an AD/ADRD Memory Study, including healthy controls. Eligibility was based on self-reported Peruvian heritage with mestizo and Quechua being the two most frequent self-descriptors. This cohort is an admixed Hispanic/Latino population with substantial Amerindian descent. Most participants were recruited in Lima and surrounding areas to the north (e.g., Huacho).

Participants were ascertained and evaluated through local memory clinics (Instituto Nacional de Ciencias Neurologicas – INCN) and community centers. Some participants were evaluated in their homes. All participants greater than 65 years of age underwent a screening and standard clinical evaluation consisting of a medical and family history interview, neuropsychological testing, behavioral and emotional assessments, and functional measures. Venous blood samples (or saliva samples when needed) were collected on all participants. All assessments were conducted in the preferred language of the participant or knowledgeable informant.

### Progressive Supranuclear Palsy (PSP)

Patients with a clinical Parkinsonism in life and neuropathological confirmation of Progressive Supranuclear Palsy (PSP) were identified from brain banks, research hospitals and neuropathologists. The top three contributing sites were the Mayo Clinic, Harvard Brain Tissue Resource Center at McClean Hospital, and the University of Pennsylvania and additional small numbers of cases were obtained from various other institutions across the US. The neuropathological diagnosis was made according to NINDS neuropathologic diagnostic criteria.  DNA was extracted from brain tissue from patients who had consented for brain donation. DNA samples and/or tissue were sent to the University of Pennsylvania for preparation for genotyping.

### Progressive Supranuclear Palsy at the University of California, Los Angeles (PSP UCLA)

This study includes subjects with Progressive Supranuclear Palsy (PSP). Subjects were enrolled in the davenutide PSP Phase Trail 2/3. Additional subjects were clinically diagnosed PSP at the University of California, San Francisco Memory and Aging Center.

Participants met the modified Neuroprotection and Natural History in Parkinson Plus Syndrome study criteria for PSP.

*See* [*https://clinicaltrials.gov/ct2/show/NCT01110720*](https://clinicaltrials.gov/ct2/show/NCT01110720) *for comprehensive inclusion and exclusion criteria.*

Boxer AL, Lang AE, Grossman M, et al. Davunetide in patients with progressive supranuclear palsy: a randomised, double-blind, placebo-controlled phase 2/3 trial. Lancet Neurol. 2014;13(7):676-685. doi:10.1016/S1474-4422(14)70088-2. PMID: [24873720](https://pubmed.ncbi.nlm.nih.gov/24873720/)

### Puerto Rican 10/66 Study (PR1066)

PR1066 ([https://www.alz.co.uk/1066/](https://nam01.safelinks.protection.outlook.com/?url=https%3A%2F%2Fwww.alz.co.uk%2F1066%2F&data=04%7C01%7CbKunkle%40med.miami.edu%7C42b90ef4de7144a46f8908d8d7342a40%7C2a144b72f23942d48c0e6f0f17c48e33%7C0%7C0%7C637495966336523289%7CUnknown%7CTWFpbGZsb3d8eyJWIjoiMC4wLjAwMDAiLCJQIjoiV2luMzIiLCJBTiI6Ik1haWwiLCJXVCI6Mn0%3D%7C1000&sdata=7EK7Sj5zKw9bT1cuYk%2FBroBoZEIw4MWdF4TNciE9M1E%3D&reserved=0)), is an Alzheimer’s Disease International study of dementia in Puerto Rico that began in 2007 (Dr. Ivonne Jimenez-Velazquez, PI). Individuals were recruited as part of the the 10/66 Population Based Study of Dementia using standard protocols. As part of this study, sociodemographic information and detailed clinical history of memory decline were collected. In addition, the Clinical Dementia Rating scale (CDR), and Community Screening Interview for Dementia, and Petersen ADL criteria were collected for all individuals. Neurocognitive testing (CERAD battery) was available for some participants. Participants were adjudicated for dementia. Lastly, the total number of samples that were whole genome sequenced is 1,565 with the breakdown of 140 Cases; 1,245 Controls and 180 MCI’s.

Prince M, Ferri CP, Acosta D, et al. The protocols for the 10/66 dementia research group population- based research programme. *BMC Public Health*. 2007;7(1):165.

### Puerto Rican Alzheimer’s Disease Initiative (PRADI)

PRADI is a NIH NIA study of late-onset Alzheimer disease focused on the Caribbean-Hispanic Puerto Rican population. Participants were ascertained for an AD/ADRD Memory Study, including healthy controls. Eligibility was based on self-reported Puerto Rican heritage. Most participants were recruited in the Island of Puerto Rico with a small fraction being ascertained in South Florida, New York, and Connecticut.

Participants were ascertained and evaluated through community centers, private memory clinics and adult day care centers. Some participants were evaluated in their homes. All participants greater than 60 years of age underwent a standard clinical evaluation consisting of a medical and family history interview, neuropsychological testing, behavioral and emotional assessments, and functional measures. Venous blood samples (or saliva samples when needed) were collected on all participants. All assessments were conducted in the preferred language of the participant or knowledgeable informant.

### Religious Orders Study/Memory and Aging Project (ROSMAP)

The Religious Orders Study (ROS) is a longitudinal, epidemiologic clinical-pathological study of memory, motor, and functional problems in older Catholic nuns, priests, and brothers aged 65 years and older from across the United States. Participants without known dementia agree to medical and psychological evaluation each year and brain donation after death. Since 1994, approximately 1,200 older persons have been enrolled and 580 are currently alive. Participants also have yearly blood draws which result in the storage of serum, plasma and cells.

The Memory and Aging Project (MAP) is a longitudinal, epidemiologic clinical-pathologic study of dementia and other chronic diseases of aging. Older persons are recruited from about 40 continuous care retirement communities and senior subsidized housing facilities around the Chicago metropolitan area. Participants without known dementia agree to annual detailed clinical evaluation and donation of brain, spinal cord and muscle after death. MAP began in 1997 and over 1,600 older adults have enrolled. Approximately 1,000 participants are currently alive. Participants also have yearly blood draws which result in the storage of serum, plasma and cells.

Clinical evaluation, self-report, and medication inspection are used to document medical conditions. The diagnostic process is the same for ROS and MAP. Briefly, a decision tree designed to mimic expert clinical judgment was implemented by computer to inform several clinical diagnoses, including dementia and AD. It combines data reduction techniques for the cognitive performance testing with a series of discrete clinical judgments made in series by a neuropsychologist and a clinician. Presumptive diagnoses of dementia and AD are calculated that conform to accepted clinical criteria. The clinician is asked to agree or disagree with the decisions. An algorithm uses these decisions to provide diagnoses of MCI and amnestic MCI. Persons with MCI are judged to have cognitive impairment by neuropsychologic testing without a diagnosis of dementia by the clinician. Persons without dementia or MCI are categorized as having no cognitive impairment (NCI).

Subjects are also evaluated neurologically every year, and, at the time of death, a review of all ante-mortem data leads to a final clinical diagnosis for each participant: each individual receives a diagnosis of syndromic Alzheimer’s disease (AD), of mild cognitive impairment (MCI), or of no cognitive impairment (NCI). After the autopsy is concluded, a spectrum of neuropathologic diagnoses are obtained, such as a pathologic diagnosis of AD as defined using the modified NIA Reagan criteria based on a modified Bielschowsky silver stain to visualize amyloid plaques and neurofibrillary tangles.

**Website:**

<https://www.rushu.rush.edu/research/departmental-research/rush-alzheimers-disease-center/radc-research/epidemologic-research>

### Research in African-American Alzheimer’s Disease Initiative (REAAADI)

REAAADI is a NIH NIA study focused on identifying genetic factors for Alzheimer disease within the African-American population in order to detect new targets for drug development and improve accessibility to Alzheimer’s disease education within the community. Participants were ascertained for multiple studies of AD/ADRD over the past 20 years, including healthy controls. Eligibility across studies was based on self-reported African American heritage. While participants were enrolled initially as part of larger studies of AD/ADRD, the first formal study focusing exclusively on African Americans began in in 2007 (Genetic Epidemiology of Alzheimer’s Disease in African Americans; AG028786). Ascertainment has continued since 2007 as part of multiple studies of AD/ADRD including the REAAADI Study (AG052410). Since 2007, participants have been ascertained via academic centers in North Carolina (Duke University, NC &T, Wake Forest University), Florida (University of Miami), New York (Columbia University), Ohio (Case Western University) and Tennessee (Vanderbilt University).

Participants were ascertained and evaluated through community centers, private memory clinics and adult day care centers. Some participants were evaluated in their homes. All participants greater than 60 years of age underwent a standard clinical evaluation consisting of a medical and family history interview,

neuropsychological testing, behavioral and emotional assessments, and functional measures. Venous blood samples (or saliva samples when needed) were collected on all participants. All assessments were conducted in the preferred language of the participant or knowledgeable informant. Note that the assessment protocols have changed over the years, but a core group of clinical measures (including neuropsychological tests) are available for all participants.

**Rotterdam Study (RS)**

The Rotterdam Elderly Study is a prospective cohort study in the Ommoord district in the city of Rotterdam, the Netherlands [Hofman et al., 1991]. Following the pilot in 1989, recruitment started in January 1990. The main objectives of the Rotterdam Study were to investigate the risk factors of cardiovascular, neurological, ophthalmological and endocrine diseases in the elderly. Up to 2008, approximately 15,000 subjects aged 45 years or over have been recruited. Participants were interviewed at home and went through an extensive set of examinations, bone mineral densitometry, including sample collections for in-depth molecular and genetic analyses. Examinations were repeated every 3-4 years in potentially changing characteristics. Participants were followed for the most common diseases in the elderly, including coronary heart disease, heart failure and stroke, Parkinson’s disease, Alzheimer’s disease and other dementias, depression and anxiety disorders, macular degeneration and glaucoma, diabetes mellitus and osteoporosis.

In the baseline and follow-up examinations participants undergo an initial screen for dementia with the Mini Mental State Examination (MMSE) and the Geriatric Mental Schedule (GMS), followed by an examination and informant interview with the Cambridge Examination for Mental Disorders of the Elderly (CAMDEX) in screen positives (MMSE <26 or GMS >0), and subsequent neurological, neuropsychological and neuroimaging examinations. Of subjects who cannot be reexamined in person, information is obtained from the GPs and the regional institute for outpatient mental health care. A consensus panel makes the final diagnoses in accordance with standard criteria (DSM-III-R criteria; NINCDS-ADRDA; NINDS-AIREN).

**Website:**

<http://www.erasmus-epidemiology.nl/research/ergo.htm>

### Stanford Extreme Phenotypes in AD (StEP AD)

This cohort contains samples from [Brains for Dementia Research](https://bdr.alzheimersresearchuk.org/) (BDR) Genetics project (Dr. Kevin Morgan, PI and Directory of the Alzheimer’s Research UK (ARUK) DNA Consortium and BDR).

The BDR cohort and program was for planned brain donation across five UK brain banks and one donation point, with standardized operating procedures, following longitudinal clinical and psychometric assessments for people with no cognitive impairment as well as those with dementia. See the Francis et al. publication for a detailed description about the BDR cohort set-up, clinical data, and psychometric assessment measures collected.

Francis PT, Costello H, Hayes GM. [Brains for Dementia Research: Evolution in a Longitudinal Brain Donation Cohort to Maximize Current and Future Value](https://www.ncbi.nlm.nih.gov/pmc/articles/PMC6294579/). *J Alzheimers Dis*. 2018;66(4):1635-1644. doi:10.3233/JAD-180699

### Texas Alzheimer’s Research and Care Consortium (TARCC)

Data from the Texas Alzheimer’s Research and Care Consortium (TARCC) includes cases enrolled at several major medical research institutions (as of 2013 this included Baylor College of Medicine, Texas Tech University Health Sciences Center, University of North Texas Health Science Center, The University of Texas Health Sciences Center at San Antonio, The University of Texas Southwestern Medical Center, and Texas A & M Health Science Center).

Individuals must be at least 55 years of age with a diagnosis of probable AD or normal cognition based on a Clinical Dementia Rating Global Score of 0. Clinical, neurological, and neuropsychological examinations performed at each site follow the TARCC research protocol that has been adopted from the standard clinical work-up for dementia. All subjects are examined at baseline and at each annual follow-up visit.

Information is obtained from the clinical and neurological examination on age at onset of symptoms (if AD patient), family history of dementia in first degree relatives, cardiovascular disease and cardiovascular disease risk factors.  Subjects also undergo a battery of neuropsychological tests as part of the TARC research protocol, with all information reviewed by a consensus panel made up of at least a physician, neuropsychologist, and research coordinator at each site to assign the final clinical diagnosis according to NINCDS-ADRDA criteria.

**Website:**

<http://www.txalzresearch.org/>

### University of Miami (MIA)

Each affected individual met NINCDS-ADRDA criteria for probable or definite AD with age at onset greater than 60 years, as determined from specific probe questions within the clinical history provided by a reliable family informant or from documentation of significant cognitive impairment in the medical record. Cognitively healthy controls were unrelated individuals from the same catchment areas and frequency matched by age and gender, and had a documented MMSE or 3MS score in the normal range.

Samples sequenced in ADSP-FUS1 include participants from the John P. Hussman Institute for Human Genomics (HIHG) Brain Bank. The HIHG Brain, Bank autopsy individuals followed the typical clinical course of disease. This sample is a clinic and community outreach sample ascertained in North and South Carolina and Virginia.

**Website:**

<http://hihg.med.miami.edu/alzheimers>

### University of Miami Brain Bank (MBB)

The University of Miami Brain Bank Cohort was ascertained through self-referred cases and controls to the [University of Miami Brain Endowment Bank](https://med.miami.edu/programs/brain-endowment-bank), a National Institutes of Health (NIH) NeuroBioBank, one of six designated brain and tissue biorepositories in the nation. Medical records are available on all cases and controls, all of whom were tested before death for cognitive function.

Subjects included in the University of Miami Brain Endowment Bank cohort include individuals who were donors to the University of Miami Brain Endowment Bank from 1986 to 2020. Donors were either individuals with history of cognitive impairment and neuropathological changes consistent with Alzheimer disease, or cognitively unaffected individuals without such neuropathological changes. Participants were recruited from community organizations, organ/tissue donation program registries, or self-referral. This is an autopsy-based study designed to analyze post-mortem brain tissue. Medical records are also obtained as available to help correlate brain behavior relationships.

**Website:**

<https://med.miami.edu/programs/brain-endowment-bank>

### University of Pittsburg (PITT)

Study participants were enrolled at the University of Pittsburgh Alzheimer’s Disease Research Center (ADRC), all of whom met the National Institute of Neurological and Communicative Disorders and Stroke/Alzheimer’s Disease and Related Disorders Association clinical criteria for probable AD. Each participant had undergone an extensive neuropsychiatric evaluation, which has been described in detail previously. Briefly, this involved a physical examination, neurological examination, semistructured psychiatric interview, and neuropsychological assessment. All patient records were reviewed at a multidisciplinary clinical consensus conference for assignment of a diagnosis. The inclusion/exclusion criteria have been published previously, but it is important to note that one of the criteria was that all the patients had to have a reliable caregiver that could provide detailed information about the patients’ clinical symptoms and their activities of daily living (ADLs). The caregiver was interviewed in person once a year and by phone every 6 months. In case of loss of a caregiver, the data were censored as the last date that reliable information was received from the caregiver.

Lopez OL, Becker JT, Saxton J, Sweet RA, Klunk W, DeKosky ST. Alteration of a clinically meaningful outcome in the natural history of Alzheimer’s disease by cholinesterase inhibition. J Am Geriatr Soc. 2005 Jan;53(1):83-7. doi: 10.1111/j.1532-5415.2005.53015.x. PMID: [15667381](https://pubmed.ncbi.nlm.nih.gov/15667381/).

### University of Washington Families (RAS)

131 families with LOAD (751 individuals) were ascertained and evaluated through the University of Washington Alzheimer Disease Research Center.  Clinical and neuropathological assessments of cases and controls, including blood sampling, medical record reviews, brain autopsies, and genetic analyses were performed under protocols approved by the institutional review boards of the University of Washington and the Seattle Veterans Affairs Puget Sound Health Care System.

### Vanderbilt University (VAN)

The UM/VU dataset contains 1,186 cases and 1,135 CNEs (new and previously published) ascertained at the University of Miami and Vanderbilt University, including 409 autopsy-confirmed cases and 136 controls. An additional 16 cases were included and 34 controls excluded from the data analyzed in the prior study. Each affected individual met NINCDS-ADRDA criteria for probably or definite AD with age at onset greater than 60 years as determined from specific probe questions within the clinical history provided by a reliable family informant or from documentation of significant cognitive impairment in the medical record. Cognitively healthy controls were unrelated individuals from the same catchment areas and frequency matched by age and gender, and had a documented MMSE or 3MS score in the normal range. Cases and controls had similar demographics: both had ages-at-onset/ages-at-exam of 74 (± 8 standard deviations), and cases were 63% female, and controls were 61% female.

### Washington Heights and Inwood Community Aging project (WHICAP)

Since inception of the study in 1992, over 6,000 participants have enrolled in the Washington Heights and Inwood Community Aging project (WHICAP). The cohort participants were nondemented initially, 65 years of age or older, and comprised of non‐Hispanic whites (32%), African Americans (28%), and Caribbean Hispanics from the Dominican Republic (44%). During each assessment, participants received a neuropsychological test battery, medical interview, and were re‐consented for sharing of genetic information and autopsy. A consensus diagnosis was derived for each participant by experienced clinicians based on NINCDS‐ADRDA criteria for possible, probable, or definite AD, or moderate or high likelihood of neuropathological criteria of AD. Every individual with whole‐exome sequencing has at least a baseline and one follow‐up assessment and examination, and for those who have died, the presence or absence of dementia was determined using a brief, validated telephone interview with participant informants: The Dementia Questionnaire (DQ) and the Telephone Interview of Cognitive Status (TICS).

Over the length of the project, WHICAP have identified environmental, health-related and genetic risk factors of disease and predictors of disease progression by collecting longitudinal data on cognitive performance, emotional health, independence in daily activities, blood pressure, anthropometric measures, cardiovascular status and selected biomarkers in this elderly, multi-ethnic cohort. Biomarker studies include lipids, amyloid peptides, sex hormones, homocysteine, insulin and C-reactive protein (CRP), and MRI in these elderly participants. WHICAP have reported that the rates of disease and the frequency of disease risk factors vary across ethnic groups, and have identified one of the largest, multi-ethnic groups of incident LOAD cases.

**Website:**

<https://www.maelstrom-research.org/mica/individual-study/whicap>

### Wisconsin Registry for Alzheimer’s Prevention (WRAP)

WRAP is an ongoing longitudinal observational cohort study of individuals age 40-65 at baseline who do not have dementia. Since 2001, WRAP has enrolled more than 1,700 individuals, 73% of whom had a parental history of probable Alzheimer’s disease (AD) dementia. Participants return for a second visit approximately 4 years after baseline, and subsequent visits occur every 2 years. At each visit, a cognitive test battery is administered, self-reported medical and lifestyle histories (e.g., diet, physical and cognitive activity, sleep, and mood) are assessed via questionnaire, and blood is drawn for laboratory tests, metabolomics, and genomics. A subset of participants have also undergone molecular imaging, structural imaging, and cerebrospinal fluid collection for biomarker measurement.

GeneRations Of WRAP (GROW) is an ancillary study to the Wisconsin Registry for Alzheimer’s Prevention (WRAP), a longitudinal cohort study enriched for individuals with a parental history of probable Alzheimer’s disease (AD) dementia. GROW is a family study that recruited extended family members of existing WRAP participants. Genomic data from family members with probable AD dementia were collected via banked brains or saliva. Individuals free of dementia and within the WRAP eligibility age of 40-65 were enrolled in WRAP. At each study visit, a cognitive test battery is administered, self-reported medical and lifestyle histories (e.g., diet, physical and cognitive activity, sleep, and mood) are assessed via questionnaire, and blood is drawn for laboratory tests, metabolomics, and genomics. A subset of participants have also undergone molecular imaging, structural imaging, and cerebrospinal fluid collection for biomarker measurement.

**Website:**

<https://wrap.wisc.edu/>

**ADSP Phenotype Harmonization Consortium Protocol**

All ADSP phenotype data are harmonized by a multi-disciplinary team that includes world experts in neuroimaging, neuropsychology, fluid biomarkers, neuropathology, and vascular contributions to ADRD. Data processing and domain-specific harmonization protocols are detailed below:

ADSP-PHC Data Processing

The Coordinating Center at Vanderbilt University Medical Center (VUMC) works directly with both existing and incoming cohort studies to curate and collate data. First, the quality of the available data is assessed, and phenotype domain(s) are assigned. If necessary, basic data cleaning or reformatting is performed by the Coordinating Center. These curated datasets are then distributed to harmonization domain teams via a secure server. Pre-statistical and statistical harmonization workflows are briefly described below and fully in each corresponding ReadMe file. Once harmonization is complete, the harmonized data files are returned to the Coordinating Center. The Coordinating Center team merges the newly harmonized phenotype data with basic demographic data and incorporates it into a larger, combined file that includes all available domain-specific harmonized data from across cohorts. Prior to sending to NIAGADS for sharing, harmonized data files are scrubbed of any PHI, including the removal of any dates, censoring age > 90 to “90+”, and subsetting to only individuals with ADSP sequencing data. The Coordinating Center at VUMC reviews and finalizes metadata produced by the harmonization teams, and files are shared with NIAGADS for release to the research community. The complete harmonized files are returned to each respective cohort study.

Cognitive Harmonization (Domain Leads: Paul Crane, MD, MPH; Jesse Mez, MD, MS)
Neuropsychological batteries differ across cohorts regarding the specific tests administered, ordering of tests, and more, complicating the ability to analyze cognitive data across cohorts. Using modern psychometrics, the Cognitive Harmonization team has developed robust methods to obtain composite scores from neuropsychological data.^1^ Briefly, a panel of content experts considers every test from each protocol and assigns it to a single primary cognitive domain (i.e., memory, executive functioning, language, visuospatial ability, or other). Tests administered consistently across cohorts serve as anchor items and facilitate co-calibration. Confirmatory factor analysis is applied to each cognitive domain, choosing between models on the basis of fit statistics and person-level impact. The output is co-calibrated scores for memory, executive functioning, language, and visuospatial functioning, which are on the same metric across cohorts and, thus, can be analyzed collectively.

Fluid Biomarker Harmonization (Domain Lead: Carlos Cruchaga, PhD)

After each incoming dataset was cleaned, Z-scores were calculated by scaling all remaining datapoints with mean 0 and variance 1 using the “scale” function in R statistical software (v3.5.2). Once the Z-scores are completed, an R script is written that inputs the incoming data file, adds the harmonized Z-scores, and outputs the harmonized biomarker dataset. Post-harmonization QC was then completed by importing the dataset into R and Excel to ensure the file is not corrupted and that formatting looks correct, and a data dictionary and readme file for each dataset was then generated.

Neuropathology Harmonization Update (Domain Leads: Gary Beecham, PhD; Tom Montine, MD, PhD)

The group first created a master data dictionary, and all incoming data summary statistics are compared to this data dictionary to ensure alignment, and any discrepancies are resolved with the contributing site. For harmonization, the incoming data dictionary is mapped with the master data dictionary, and identify three types of variables: (1) those that need to be recorded, (2) those that need to be derived, and (3) those that passed through without modification needed. Once mapping is complete, an R script is written that outputs the harmonized neuropathology dataset. Post-harmonization QC included the generation of summary statistics of the harmonized dataset and the import of the harmonized dataset into R and Excel to ensure that it is not corrupted and that formatting looks correct.

Cardiovascular Risk Factor Harmonization (Domain Leads: Richard Mayeux, MD, MSc; Adam Brickman, PhD)

Self-reported data from each participant were recorded as a binary indicator (Yes-Ever Had or No-Never Had) for heart disease, hypertension, and diabetes.^2^ Details on heart conditions varied between groups, so report of any heart disease qualified as presence of heart disease in this study. A quantitative variable was recorded for body mass index (BMI) from the last visit. The PCAMix package computes principal components (PCs) in a mixture of quantitative and qualitative data and was used to summarize the cardiovascular risk factors into one summary score by computing PCs from the four vascular variables. The goal of the dimensionality reduction was to capture the greatest amount of variance accounted for by the four vascular risk factors, and thus each participant’s values from the first principal component from each of the cohorts was used as their vascular risk factor score (CVRscore). 
 

Structural MRI Harmonization (Domain Leads: Shannon Risacher, PhD; Christos Davatzikos, PhD)

Original files in DICOM format were converted to NIFTI format using the dcm2niix conversion tool^3^ or Freesurfer’s mri_convert tool. MRI images in NIFTI format and associated json files with detailed scanner information were the consolidated into the final image dataset. Data consolidation included automated and semi-automated steps for identification and selection of the T1 scan at each timepoint, visit, quality control of images, standardization of image names, and extraction of meta-data with scanner information from individual json files and LONI download information. Preprocessing, anatomical segmentation, and harmonization were then completed for both Freesurfer and MUSE pipelines^4^, and these processes are further detailed in ADSP-PHC-T1_README_2023.12.01.docx, available via NIAGADS.

PET Harmonization (Domain Leads: Elizabeth Mormino, PhD: Duygu Tosun-Turgut, PhD)

The MRI-free amyloid (Aβ) PET workflow follows the pipeline outlined by Laundau et al.^5^ Tracer-specific SUVR-to-CL equations were derived for the MRI-Free pipeline using the level 2 GAAIN CL analysis method. Next, a data-driven Gaussian Mixture Model (GMM) approach was used to extract subject-level probabilities reflecting membership to amyloid positive (A+) or negative (A-) clusters within each amyloid PET dataset.^6^ The MRI-free tau PET pipeline was built using the same amyloid PET MRI-free PET processing pipeline^5^ but adapted to include multiple templates during the non-linear warping stage, in contrast to the amyloid MRI-free pipeline which uses a single universal template. Quality control procedures were divided into two categories: (1) QC Timing, and (2) QC Image. The complete harmonization workflow is detailed in ADSP-PHC-PET-Scalar_WhitePaper_2023.12.01.docx, available via NIAGADS.

Diffusion MRI Harmonization (Domain Leads: Bennett Landman, PhD; Derek Archer, PhD)

Following the organization of raw data BIDS structure, data were processed via either the PreQual^7^ or SLANT-TICV^8^ pipelines. The resulting tensor maps were then used to calculate the diffusion scalar maps for fractional anisotropy (FA), mean diffusivity (MD), radial diffusivity (RD), and axial diffusivity (AD) with MRTrix3’s *tensor2scalar^9^.* The outputs from each pipeline are kept in the original formatting after preprocessing was run. An additional quality assurance check was performed on the SLANT-TICV segmentation and Prequels preprocessing output images, as well as the scalar metrics obtained from tractseg and the DWI scalars for each ROI in the Eve Type 3 atlas. The complete harmonization workflow is detailed in ADSP-PHC-DTI_README_2023.12.01.docx, available via NIAGADS.

FLAIR Harmonization (Domain Leads: Adam Brickman, PhD; Mohamad Habes, PhD)

FLAIR DICOM files were converted to NIfTI format using FreeSurfer’s standard converter, *mri_convert.* The images were conformed to voxel dimesnsions of 1x1x1 mm^3^ and spatial dimensions of 256x256x256 using the *–conform* argument in *mri_convert*. Subsequently, FLAIR images underwent bias field correction and intensity normalization to a range of 0-255. This was accomplished using *nu_correct.mni* from the Montreal Neurological Institute, which employs N3 Bias Field Correction—a method more suitable for FLAIR images than N4. A percentile threshold was calculated to distinguish between the bright and brightest voxels in the FLAIR image based on its contrast-to-noise (CNR) ratio. The images then underwent segmentation using a Gaussian Mixture Model and the Expectation-Maximization algorithm. The model was initiated using the calculated percentile threshold. Finally, FreeSurfer’s mri_segstats tools were used to compute the volume.
